# Human RIG-I Antiviral Deficiency Caused by a Dominant-Negative Variant Locked in a Signaling-Inactive State

**DOI:** 10.64898/2026.03.02.26347088

**Authors:** Mihai Solotchi, Huie Jing, Emma Gebauer, Scott J Novick, Bruce D Pascal, Wesley Tung, Pranita Hanpude, Talia Madura, Yu Zhang, Camille Alba, Annalisa Saracino, Paola Laghetti, Elana R Shaw, Lindsey B Rosen, Steven M Holland, Andrea Lisco, Clifton L Dalgard, Joseph Marcotrigiano, Patrick R Griffin, Helen C Su, Smita S Patel

## Abstract

RIG-I is a cytosolic immune receptor that provides the first line of defense by detecting viral RNA and triggering antiviral responses. Its physiological role in humans remains unclear, as no patients with complete RIG-I deficiency have yet been reported. We identified a critically ill COVID-19 patient with severe RIG-I deficiency caused by heterozygous RIG-I G731R, a novel dominant loss-of-function variant. The G731R mutation in helicase motif VI disrupts the arginine finger, impairing the ATPase activity of RIG-I, but not its RNA-binding ability. However, viral RNA binding fails to expose the signaling domains, thereby impairing the IFN-β response of G731R. Instead, G731R competes with wild-type RIG-I, exerting a dominant negative effect. The loss-of-function is caused by bulky-charged substitutions at G731, as alanine or leucine substitution results in an unexpected gain-of-function phenotype. These findings highlight the importance of uncompromised RIG-I function for human antiviral immunity and the pleiotropic effects of single mutations.

## INTRODUCTION

Viral infections present a constant threat to human health, necessitating a swift and precise host innate immune response to effectively limit viral growth and disease progression. Central to this response are the RIG-I-like nucleic acid receptors (RLRs), a family of cytosolic pattern recognition receptors, which surveil the cell for signs of viral RNA^1–5^. The RLRs are comprised of three proteins, RIG-I (Retinoic acid-inducible gene I, *RIG-I*), MDA5 (Melanoma differentiation-associated gene 5, *IFIH1*), and LGP2 (Laboratory of Genetics and Physiology 2, *DHX58*) that belong to the DExH/D-box Superfamily 2 (SF2) RNA helicases. The RLRs rapidly identify the virus and trigger a cascade of signaling events that induce the expression of type I interferons (IFNs) that activate the antiviral genes to fight the infection^6–9^.

Among the RLRs, RIG-I plays a frontline role in initiating an antiviral response by recognizing and binding to viral RNAs that contain a 5’ triphosphate (5’ppp) at a blunt double-stranded (ds) RNA end^10–13^. RIG-I is a multidomain protein with a centrally located recA-like helicase domain (Hel1, Hel2), a nested insertion domain (Hel2i), a C-terminal domain (CTD) that recognizes the 5’ppp RNA-end, and N-terminal tandem caspase activation and recruitment domains (CARDs) serving as the signaling domains ^14–16^ (Figure 1A). In the absence of RNA, RIG-I assumes an autoinhibited conformation wherein the CARDs are sequestered by Hel2i through a CARD2:Hel2i interface ^16,17^ stabilized by a ∼60 amino acid intrinsically disordered CARDs-Helicase linker (CHL - dotted lines in Figure 1A, 1B). RNA binding disrupts the interface and releases the CARDs for activating the signaling pathway by partnering with the Mitochondrial Antiviral Signaling protein (MAVS). RNA binding also activates the ATPase activity of RIG-I, which serves as a proofreading mechanism to distinguish viral RNAs from host RNAs^16,18^. RIG-I’s ATPase activity drives its movement along RNA molecules, which promotes rapid dissociation from host RNAs that lack a 5’ppp. However, when RIG-I encounters viral RNA, specific interactions with the 5’ppp slow this translocation. The prolonged interaction with 5’ppp RNA allows RIG-I’s CARD domains to engage with MAVS, triggering interferon and antiviral response^19–21^. In Singleton-Merten syndrome (SMS), characterized by cardiovascular, dental, and skeletal abnormalities but without increased susceptibility to virus infections^22^, ATPase-deficient RIG-I mutants form unusually stable complexes with host RNAs, constitutively activating RIG-I signaling^18^.

**Figure 1.**
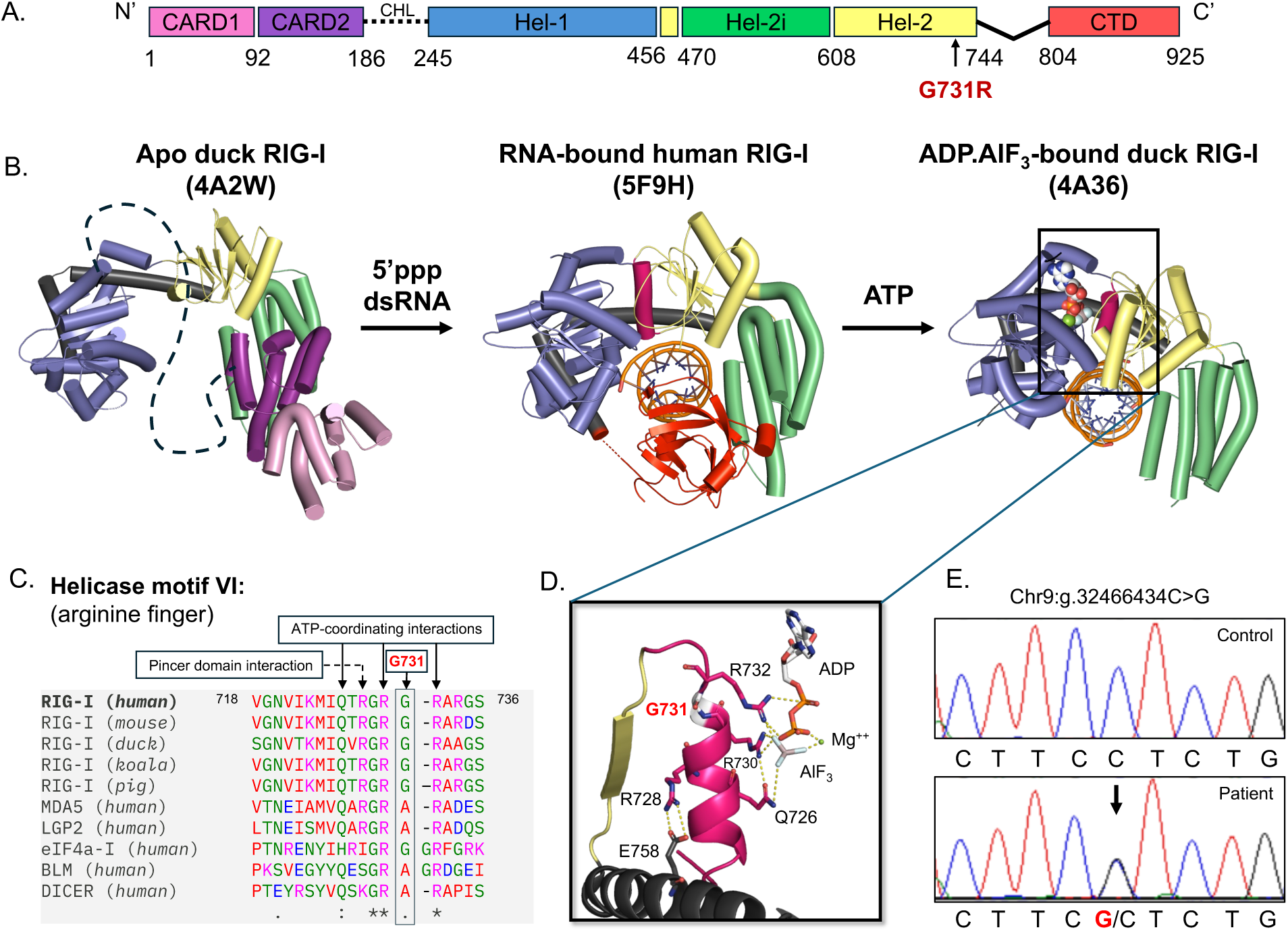
**Novel RIG-I G731R mutation identified in a severe COVID-19 patient.** (A) Domain structure of human RIG-I. The arrow indicates the location of the G731R mutation. (B) RNA- and ATP-induced conformational activation of RIG-I. Top view of apo duck RIG-I (PDB: 4A2W) with CARDs (pink/purple) sequestered to Hel2i domain (green) autoinhibitory interface and helicase domains in an open conformation, semi-closed 5’ppp RNA-bound human RIG-I (PDB: 5F9H), and closed RNA-bound duck RIG-I with ADP·AlF_3_ with missing CTD (PDB: 4A36). (C) Clustal Omega Multiple Sequence Alignment^54^ of helicase motif VI in RIG-I of various species, RLRs, and other SF2 RNA helicases. (D) Zoomed-in view of motif VI (in pink) in the closed state of RIG-I and the associated interactions with ATP and the pincer domain helix (black). (E) Chromatograms from Sanger sequencing of genomic DNA of the patient showing a heterozygous C to G mutation indicated by the arrow.

Patients with inborn errors of immunity have demonstrated the importance of genes regulating the type I IFN pathway for antiviral immunity in humans^23,24^. Partial defects in RIG-I resulting from haploinsufficiency or a common cis-expression Quantitative Trait Locus (eQTL) have been linked to severe influenza or acute Hepatitis E virus infections^25–27^. However, the full physiological role of RIG-I in humans remains incompletely understood, as no patients with complete RIG-I functional deficiency have yet been reported. Heterozygous loss-of-function or hypomorphic RIG-I mutants do naturally occur in humans, but severe RIG-I deficiency caused by variants that confer <50% of wild-type (WT) activity has not been identified^28^. RIG-I knockout (KO) mice exhibit embryonic lethality due to massive liver apoptosis ^29,30^ or spontaneously developed colitis-like symptoms ^31^ and progressive myeloproliferative disorders^32^, suggesting that RIG-I may play an important role in human development or immune homeostasis in addition to functioning as an antiviral pattern recognition receptor.

Recently, studies have highlighted certain roles of RIG-I in response to SARS-CoV-2, including abrogation of the early viral replication stages ^33^ and prophylactic protection of mice from lethal viral infection^34^. Additionally, SARS-CoV-2 has several mechanisms in place to hinder RIG-I signaling directly and indirectly^35,36^. Here, we identified a novel RIG-I G731R variant in a patient with critical respiratory COVID-19. This G731R variant has not been previously reported in genome databases, and the mechanism of action causing its pathogenicity remains elusive. The G731R mutation is located in helicase motif VI, which is conserved in RLRs and SF2 family helicases ^37^ (Figure 1C). Most notably, the arginine fingers flanking G731 interact with ATP, promoting ATP hydrolysis (Figure 1D).

To understand how the G731R mutation disrupts RIG-I activation, we used a combination of cellular reporter assays, biophysical and biochemical functional assays, and hydrogen-deuterium exchange mass spectrometry (HDX-MS) structural studies.

Our results show that the G731R mutant binds normally to 5’ppp RNA ligand, but is deficient in ATPase activity and fails to initiate downstream signaling events. Critically, G731R acts as a competitive inhibitor against WT RIG-I. This makes RIG-I G731R the first naturally occurring dominant-negative RIG-I variant identified in a human, resulting in severe functional RIG-I deficiency. Mechanistically, G731R adopts a CARDs-protected conformation on 5’ppp RNA that is kinetically unstable and resembles the previously described “CTD-mode” conformation of RIG-I^18^. The mutation effectively locks RIG-I in a signaling-inactive intermediate state. This unexpected loss-of-function mechanism provides valuable insights into RIG-I activation, which can inform the design of therapeutic RIG-I modulators targeting specific helicase motifs.

## RESULTS

### Identification of RIG-I G731R variant in a patient with life-threatening COVID-19

Since numerous studies using mouse models and tissue culture systems support a role for RIG-I in antiviral immunity^29,30,38,39^, we hypothesized that pathogenic variants associated with impaired RIG-I function might exist in patients with severe virus infections. In our longstanding search among patients referred to us for increased susceptibility to various respiratory viruses, including SARS-CoV-2 and influenza virus, we identified one such patient who was hospitalized in early Spring of 2020 with critical COVID-19 pneumonia. The patient, a male in his early 60s, had neither comorbidities, nor previous history of severe viral infections, nor any family history of virus-associated diseases. He presented with a nine-day history of worsening fever, cough, and dyspnea. Radiological evaluation in the emergency department revealed diffuse lung infiltrates, and a nasopharyngeal swab molecular assay was PCR positive for SARS-CoV-2. Despite treatment with azithromycin, hydroxychloroquine, and lopinavir/ritonavir, he required invasive mechanical ventilation for acute respiratory distress, as well as therapeutic anticoagulation with low molecular weight heparin (LMWH) for incidental thromboembolic complications. After ten days in the intensive care unit (ICU), he was transferred to the inpatient service and stayed for a total of 34 days before hospital discharge. Subsequently, he received influenza and COVID-19 immunizations and has not developed any new infectious complications.

During his hospitalization, the patient had elevated CRP (147 mg/L), unremarkable numbers of polymorphonuclear leukocytes (5.305 x 10^9^/L, normal range 4.0-11.3 x 10^9^/L), and decreased lymphocyte numbers (0.449 x 10^9^/L, normal range 1.320-3.600 x 10^9^/L). Autoantibodies against type I IFNs (IFN-α2, IFN-β, or IFN-ω), which can neutralize endogenously produced type I IFNs to increase COVID-19 disease severity^40,41^, were not detected in the patient’s plasma (106, 254, 326 fluorescence intensities, well below the normal cutoffs of <1,310, <386, and <1,387 fluorescence intensities for anti-IFN-α2, anti-IFN-β, and anti-IFN-ω autoantibodies, respectively).

Whole genome sequencing revealed a heterozygous *RIGI* variant (chr9:g.32466434C>G, NM_014314.4:c.2191G>C, NP_055129.2, p.Gly731Arg), which was confirmed by Sanger dideoxy sequencing (Figure 1E). No variants were detected in other genes known to regulate the type I IFN pathway (Table S1). This *RIGI* variant has not been previously reported in the Genome Aggregation Database (gnomAD) population database and was computationally predicted to be deleterious (CADD score 29.7 and AlphaMissense score 0.9725).

### The G731R loss-of-function RIG-I variant exerts dominant negative (DN) effects

To evaluate whether the G731R variant alters RIG-I protein function, the IFN response was tested using a dual luciferase IFN-β promoter-reporter assay in HEK293T RIG-I^-/-^cells. Cells were transfected with either an empty vector (EV) or with a plasmid containing RIG-I (WT or G731R variant) under a constitutively overexpressing cytomegalovirus (CMV) promoter. The RIG-I-dependent IFN response was quantitated as the luminescent activity of Firefly relative to Renilla (STAR methods). Without RNA stimulation, cells transfected with WT RIG-I displayed a low basal IFN-β response, but when challenged with a prototypical RIG-I ligand, 5’ppp ds39 RNA^5,42–44^, RIG-I robustly stimulated the IFN-β promoter response (Figure 2A). By contrast, the G731R variant was poorly stimulated by the 5’ppp ds39 RNA transfections, even at maximal concentrations of ligand.

**Figure 2.**
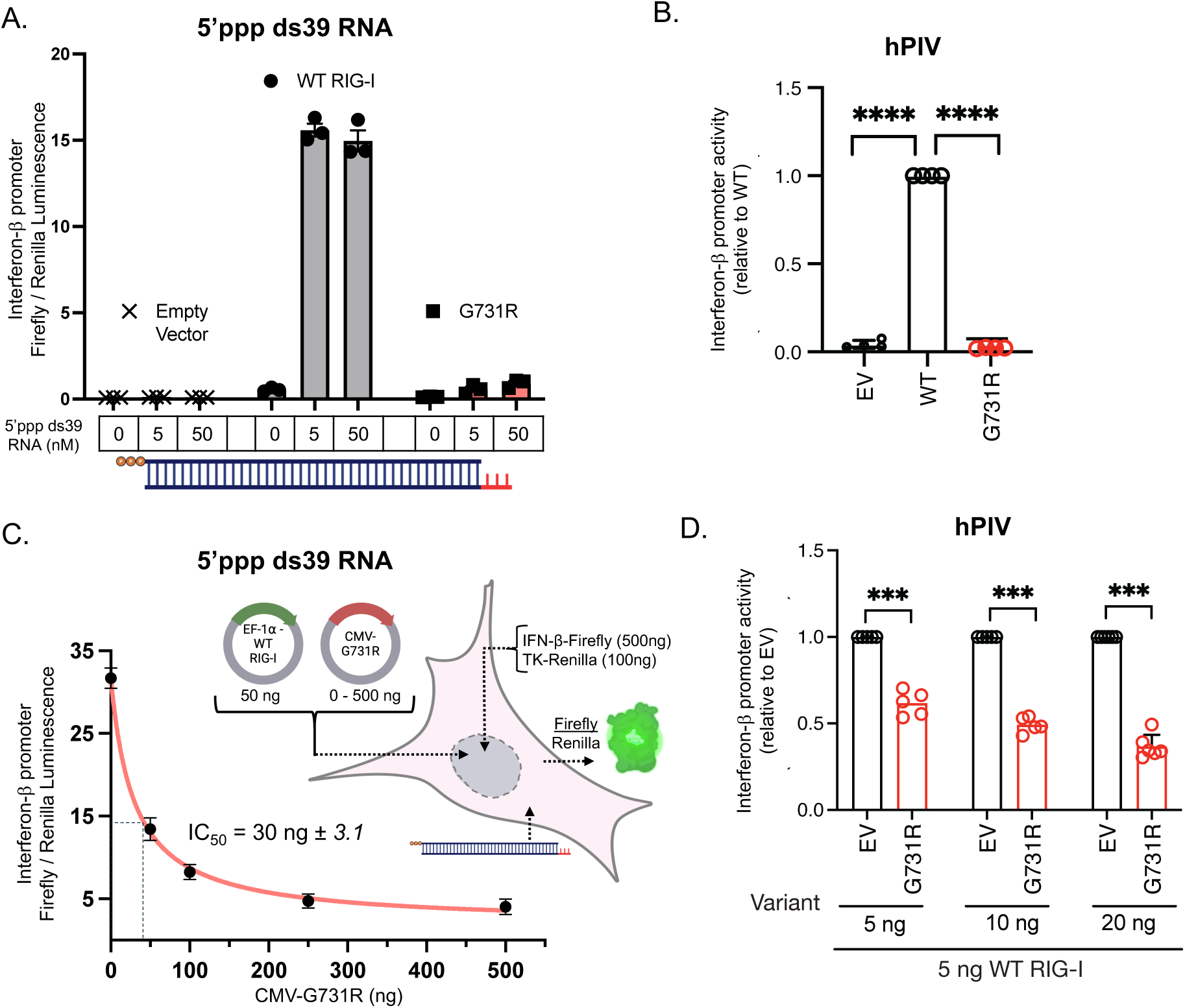
**RIG-I G731R is a loss-of-function variant with a dominant negative effect.** (A) 5’ppp ds39 RNA induced IFN-β reporter activities after transfection with empty vector (EV), WT RIG-I, and G731R plasmids (50 ng) in HEK293T RIG-I^-/-^ cells (Star Methods) (technical replicates, n=3). (B) hPIV-induced IFN-β reporter luciferase activities for G731R. HEK293T cells were transfected with either the WT or G731R construct or EV along with the IFN-β-Luc reporter and a constitutively expressed Renilla reporter for 6 hours. Cells were then infected with hPIV (MOI 3) for an additional 20 hours before measuring luciferase activity. IFN-β-Luc reporter activity in each condition was first normalized to Renilla and then presented relative to WT RIG-I. Data show means ± SD from 3 independent experiments. Statistical analysis was performed using one-sample t-test. ****, *P*<0.0001. (C) IFN-β promoter response measured by dual luciferase reporter assay in HEK293T RIG-I^-/-^ cells transfected with a constant amount of WT RIG-I plasmid (50 ng), increasing amount of G731R plasmid (0-500 ng), and 5 nM 5’ppp ds39 RNA. The error bars represent the standard error of the mean (technical replicates, n=3), and the curve represents fitting of the mean data to equation 1 to obtain the half maximal inhibitory concentration IC_50_. (D) hPIV-induced IFN-β reporter luciferase activities in cells co-transfected with WT RIG-I (5 ng) and increasing doses (5 to 20 ng) of G731R or EV plasmids. IFN-β-Luc reporter activity in each condition was first normalized to Renilla and then shown relative to EV. Data show means ± SD from 3 independent experiments. Statistical analysis was performed using one-sample t-test. ***, *P*<0.001

Furthermore, RIG-I has been shown to be essential for interferon production in response to RNA viruses, including paramyxoviruses^30^. Infection with human parainfluenza virus (hPIV), a member of the paramyxovirus family, induced robust luciferase activity in cells overexpressing WT RIG-I (Figure 2B). By contrast, no luciferase activity was observed in cells overexpressing the G731R variant (Figure 2B). Western blots confirmed similar expression levels of the RIG-I proteins (Figure S1A, S1B, S1C). Therefore, we establish that RIG-I G731R is a naturally occurring loss-of-function (LOF) RIG-I variant.

As the G731R variant was heterozygous in the patient, we investigated whether this variant might exert a dominant negative effect. WT RIG-I was co-transfected with increasing amounts of G731R, together with the luciferase reporter plasmids. G731R decreased the IFN-β promoter response of WT RIG-I in a dose-dependent manner in the presence of 5’ppp ds39 RNA (Figure 2C, S1B) but not in the absence of RNA (Figure S1D). Similarly, G731R decreased hPIV induced IFN-β reporter activity of WT RIG-I in a dose-dependent manner (Figure 2D). These findings demonstrate that G731R is a dominant negative loss-of-function RIG-I mutant, suggesting that it may severely compromise RIG-I activity in the heterozygous patient (G731R/WT).

### G731R binds to 5’ppp dsRNA with a high affinity

As this patient is the first reported to display near complete RIG-I functional deficiency, we sought to understand the biochemical mechanism underlying the dominant-negative effect of the G731R mutation. We first assessed whether the mutant can engage with 5’ppp dsRNA. Electrophoretic Mobility Shift Assay (EMSA) was used to test the binding of purified RIG-I protein to the 5’ppp ds39 RNA, the ligand that activated WT RIG-I in reporter assays (Figure 2A). With a footprint of about 10 bp, the ds39 RNA can accommodate multiple RIG-I molecules. However, RIG-I has a high affinity for the 5’ppp RNA end, but in the presence of ATP, RIG-I can translocate internally, and multiple

RIG-I molecules can bind to the long RNA^18,43^. Accordingly, WT RIG-I bound to 5’ppp ds39 RNA as monomers and dimers and transitioned to form higher oligomers in the presence of ATP (Figure 3A). Interestingly, G731R was not defective in RNA binding, but multimeric RIG-I complexes were reduced on the 5’ppp ds39 RNA with or without the addition of ATP. This demonstrates that G731R binds to 5’ppp RNA, but it does so primarily as a monomer.

**Figure 3.**
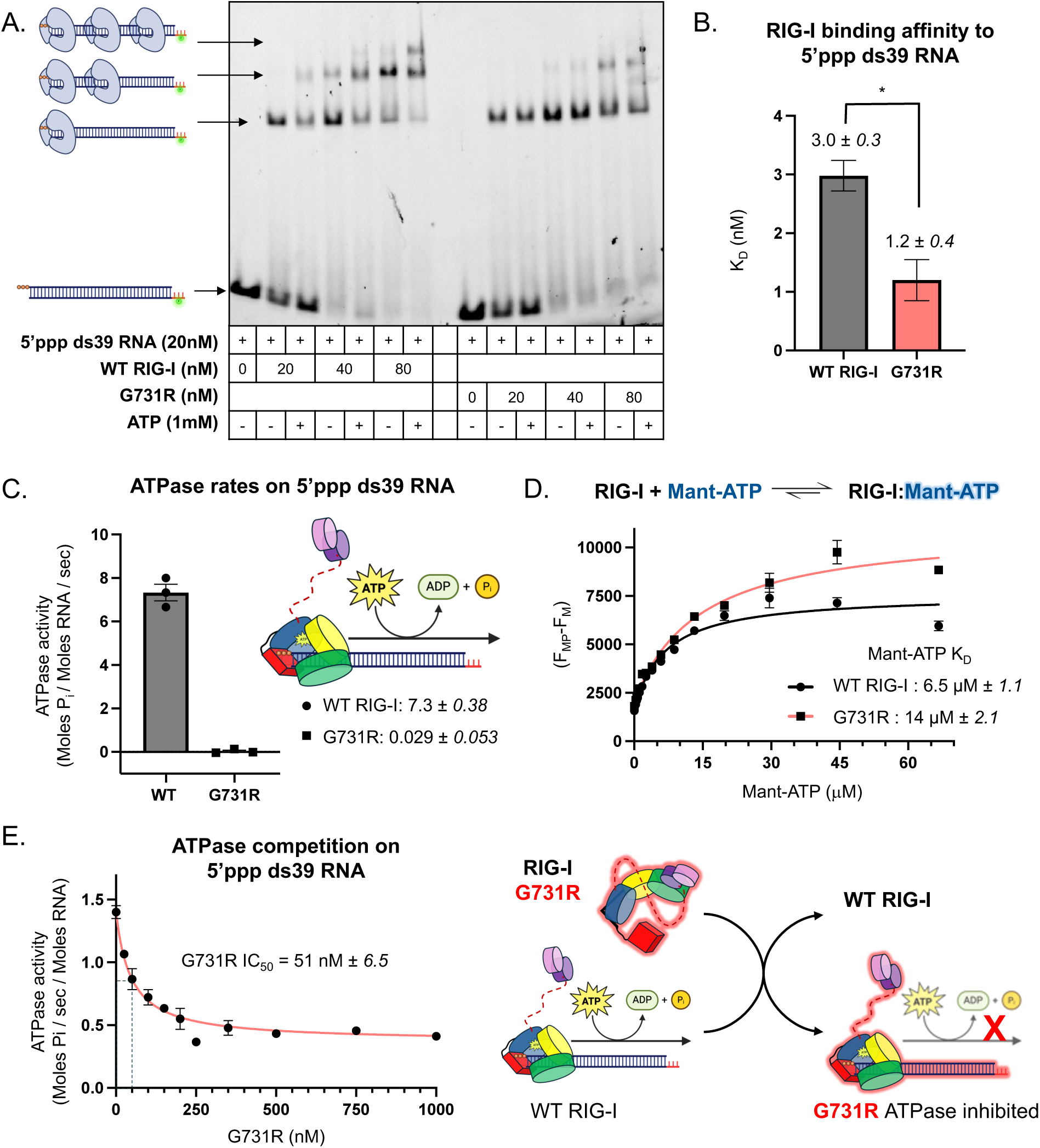
**RIG-I G731R competitively binds 5’ppp dsRNA in an ATPase-deficient manner.** (A) Electrophoretic mobility shift assay of fluorescently labeled 5’ppp ds39 RNA upon addition of WT and G731R RIG-I (representative image from n=3). (B) 5’ppp ds39 RNA binding affinity (K_D_) was obtained using a competitive fluorescence-based RNA binding assay (Figure S2, Star Methods). Data are mean ± SE from 3 independent experiments. Statistical analysis was performed using unpaired t-test. *, P<0.05. (C) ATPase rates of WT and G731R RIG-I at 1 µM concentration were measured in the presence of 5 nM 5’ppp ds39 RNA. Mean rates of hydrolysis (Moles P_i_ / Moles RNA / sec) are reported (technical replicates, n=3). (D) Mant-ATP binding to WT and G731R RIG-I using fluorescence-based titration. The differences in the fluorescence intensity of Mant-ATP alone (F_M_) and Mant-ATP with protein (F_MP_) are plotted against Mant-ATP concentration. The error bars represent the standard error of the mean (n=3), and the curve represents hyperbolic fitting of the mean data to **equation 2** (Star Methods) to obtain the indicated Mant-ATP K_D_ values. (E) G731R inhibits the RNA-dependent ATPase activity of WT RIG-I in a dose-dependent manner. ATPase activity was measured in the presence of 5’ppp ds39 RNA at constant WT RIG-I and increasing G731R protein. The error bars represent the standard error of the mean (n=3), and the curve represents fitting of the mean data to **equation 1** (Star Methods) to obtain the indicated IC_50_ value.

To determine the equilibrium binding affinity of G731R for the 5′ppp ds39 RNA, we performed a fluorescence polarization-based RNA competition assay. In this assay, WT or G731R bound to fluorescein-labeled 5′ppp 10-bp hairpin (HP) RNA was challenged with increasing concentrations of unlabeled 5′ppp ds39 RNA (Figure S2). The resulting competition curves yielded K_D_ values of 3 nM for WT and 1.2 nM for G731R (Figure 3B; Star Methods; Figure S2), indicating that G731R binds 5′ppp ds39 RNA with ∼3-fold higher affinity than WT RIG-I (P = 0.0151). This enhanced affinity, combined with its loss-of-function phenotype, may explain the dominant-negative behavior of G731R. Notably, the G731R complex with 5′ppp HP RNA exhibited a lower fluorescence polarization amplitude compared to WT (Figure S2C), suggesting conformational differences between WT and G731R in the RNA-bound state.

### G731R is ATPase-deficient and competes with WT RIG-I for 5’ppp RNA binding

The G731 residue lies within helicase motif VI, positioned between two critical arginine residues that coordinate ATP phosphates during catalysis (Figure 1C,D). Substitution of G731 with a bulky, positively charged arginine could perturb ATP binding or hydrolysis by altering the local structure or charge environment. To test this, we measured ATPase activity. Because RIG-I hydrolyzes ATP only in the presence of RNA, assays were performed with 5′ppp ds39 RNA. WT RIG-I exhibited robust RNA-dependent ATP hydrolysis with a rate constant of 7.3 s⁻¹, whereas the G731R mutant showed severely impaired activity, with a rate constant of only 0.03 s⁻¹ (Figure 3C). To assess whether this defect arose from impaired ATP binding, we measured binding affinity using fluorescent Mant-ATP. G731R bound Mant-ATP with a K_D_ of 14 μM, compared to 6.5 μM for WT RIG-I (P=0.0061) (Figure 3D). However, this ∼2-fold difference is unlikely to be physiologically relevant given the intracellular ATP concentration of ∼3 mM^45^. Thus, while G731R can bind ATP, it is clearly defective in ATP hydrolysis.

Furthermore, increasing concentrations of G731R decreased the 5′ppp ds39 stimulated ATPase activity of WT RIG-I (Figure 3E). Since RIG-I hydrolyzes ATP only in the presence of RNA, these results demonstrate that G731R competes with WT RIG-I for 5′ppp RNA binding, thereby explaining its dominant-negative effect when coexpressed with WT. Notably, even at 1 μM G731R, 20-fold higher than WT RIG-I, the ATPase activity of WT RIG-I was not fully inhibited. We suspect that this residual activity reflects WT RIG-I molecules bound to the stem region of 5′ppp ds39 RNA within mixed G731R–WT complexes. Such stem-bound RIG-I complexes are kinetically transient and unlikely to contribute to downstream signaling activity^18,43^. Collectively, these results indicate that G731R engages 5′ppp RNA with high affinity, competing with WT RIG-I for RNA binding. Because the G731R–RNA complex is signaling-inactive, it suppresses the antiviral RNA immune response mediated by WT RIG-I.

### The G731R mutation prevents exposure of the CARDs-CHL upon RNA binding

The loss-of-function phenotype of G731R that binds 5’ppp RNA and ATP cannot be due to its ATPase deficiency, because SMS variants, such as C268F and E373A, that are ATPase-inactive exhibit gain-of-function phenotypes^46,47^. One possible explanation is that G731R binds 5’ppp RNA in an altered conformation that keeps the CARD domains autoinhibited. To test this hypothesis, we used hydrogen-deuterium exchange coupled to mass spectrometry (HDX-MS), a technique previously used to assay CARDs exposure upon RNA binding^43,48–50^. HDX-MS measures the time-dependent exchange of backbone amide hydrogens with deuterium, detected by mass spectrometry. The rate of exchange reflects local protein structure: rapid exchange corresponds to unfolded regions, flexible loops, or solvent-exposed areas, whereas slow exchange indicates stably folded or buried regions. We present the HDX-MS results as time-consolidated differential patterns to compare conformational states of RIG-I under different conditions (e.g., with or without ligand, or between WT and mutant).

Differential HDX-MS showed that in the absence of RNA, there is decreased solvent exchange in the CARD1 domain and the CHL of G731R relative to WT RIG-I (Figures 4A,B, S3–S5). Furthermore, increased mild protection throughout the helicase domains and CTD of G731R, including helicase motif IVa (aa 661–681) was observed in G731R relative to WT. Thus, G731R in the absence of RNA adopts a slightly more solvent-protected conformation than WT RIG-I.

**Figure 4.**
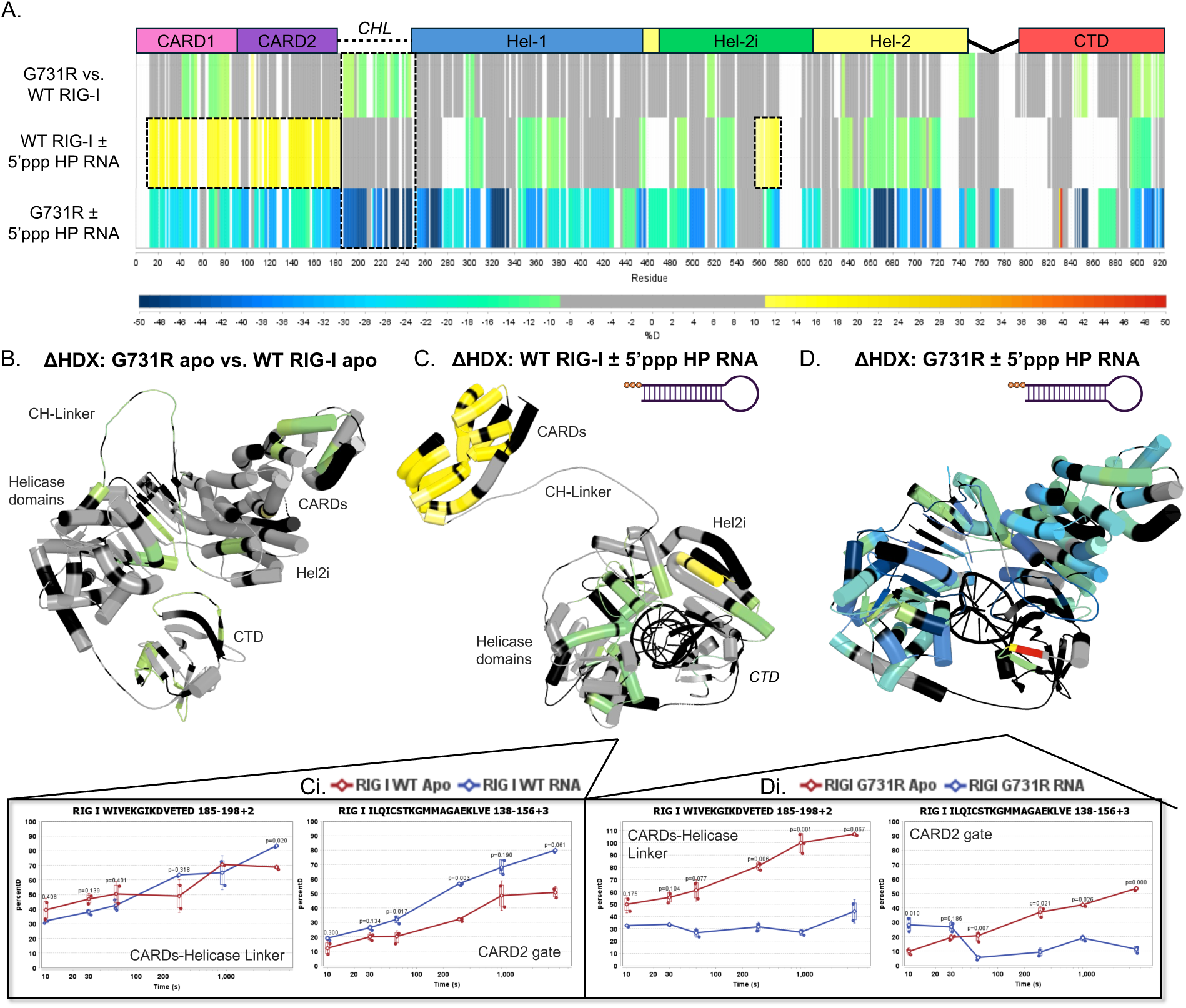
**HDX-MS analysis of G731R reveals CARDs-CHL protection.** (A) Linear mapping of the HDX differential data of WT and G731R with and without 5’ppp HP RNA. In the colorimetric HDX scheme, shifting towards red indicates a higher degree of solvent exchange, while blue indicates less. If there is no significant difference in the solvent exchange of two samples analyzed side-by-side, the HDX differential remains gray. The white/black areas indicate that the peptides were not resolved for mass spectrometry. (B) The differential HDX pattern of G731R apo vs WT RIG-I apo mapped onto a 3D composite model of G731R apo (4A2W, 3TBK, 4AY2, & computationally modeled CH-Linker). (C) The differential HDX pattern of WT RIG-I ± 5’ppp HP RNA mapped onto a 3D composite model of RNA-bound WT RIG-I with computationally modeled CHL and CARDs in the exposed state (PDB: 5E3H, 4P4H, 4AY2). (Ci) Deuterium uptake plots of select RIG-I peptides represented as percent deuterium uptake over time. (D) The differential HDX pattern of G731R ± 5’ppp 10bp HP RNA mapped onto a composite model of RNA-bound G731R with computationally modeled CHL and CARDs sequestered by Hel2i (PDB: 4A2W, 3TBK, 4AY2). (Di) Deuterium uptake plots of select RIG-I peptides represented as percent deuterium uptake over time.

HDX-MS of WT RIG-I with and without 5’ppp 10-bp hairpin RNA (Figures 4A,C, S6,7) revealed patterns characteristic of RNA engagement and CARD domain exposure. The CARD domains (aa 1-186) and the CARD2:Hel2i interface, comprised by the CARD2-gate region (aa 138-156) (Figure 4C-i) and the Hel2i region (aa 556-578), showed increased solvent exchange. Thus, as expected, 5’ppp RNA binding disrupts the CARD2:Hel2i interface of WT RIG-I and leads to exposure of the CARD domains.

Another small region of the Hel2i domain (aa 520-538), also part of the CARD2:Hel2i interface, exhibited protection upon RNA binding, consistent with this Hel2i region forming a junction with the CTD (aa 895-915)^15,16,51^. Finally, we observed protection patterns throughout the Hel1 and Hel2 domains, coinciding with regions known to interact with the RNA backbone. Overall, these HDX patterns are consistent with RNA-induced conformational changes that lead to CARD domain exposure in WT RIG-I.

The HDX-MS pattern of G731R in the presence and absence of RNA differed significantly from that of WT RIG-I. In contrast to the increased solvent exchange observed in the CARD domains of WT RIG-I upon RNA binding, G731R showed an overall protection of the CARDs and the CHL region (Figures 4A,D, S8,9). The helicase domains showed protection consistent with RNA binding but a distinct pattern from WT RIG-I. Regions showing distinctly high protection included helicase motif IVa and the Hel2 loop (aa 661-693), as well as the capping loop (aa 842-855) in the CTD that interacts with the 5’ppp RNA end. Overall, the HDX-MS suggests that the CARDs and CHL regions are protected from solvent exchange in the G731R, which is consistent with its impaired IFN-β response. Furthermore, the distinct protection pattern within the helicase domain indicates that G731R binds RNA in a conformation that differs substantially from the WT protein.

### A small deletion in the CHL partially rescues the activities of G731R

The protection of the CHL region in G731R in the presence of RNA was surprising. Previous studies have shown that CHL (aa 183-245) (Figure 5A), a negatively charged, intrinsically disordered region, stabilizes the autoinhibited conformation of RIG-I, and 10 aa deletions within the CHL disrupt this regulation and increase the basal signaling response of RIG-I^43^. We wondered if disrupting the CHL would rescue the activity of G731R. For this, we used the well-characterized ι1190-200 mutant. Consistent with previous findings, the ι1190-200 RIG-I showed a ∼10-fold higher basal IFN-β signaling response than full-length WT RIG-I and a 2-fold higher 5’ppp ds39 RNA-stimulated response (Figure 5B). Interestingly, the 190-200 deletion in G731R increased both the basal and the 5’ppp RNA IFN-β response by 10-fold (Figure 5B, S1E), thus partially rescuing the RNA response.

**Figure 5.**
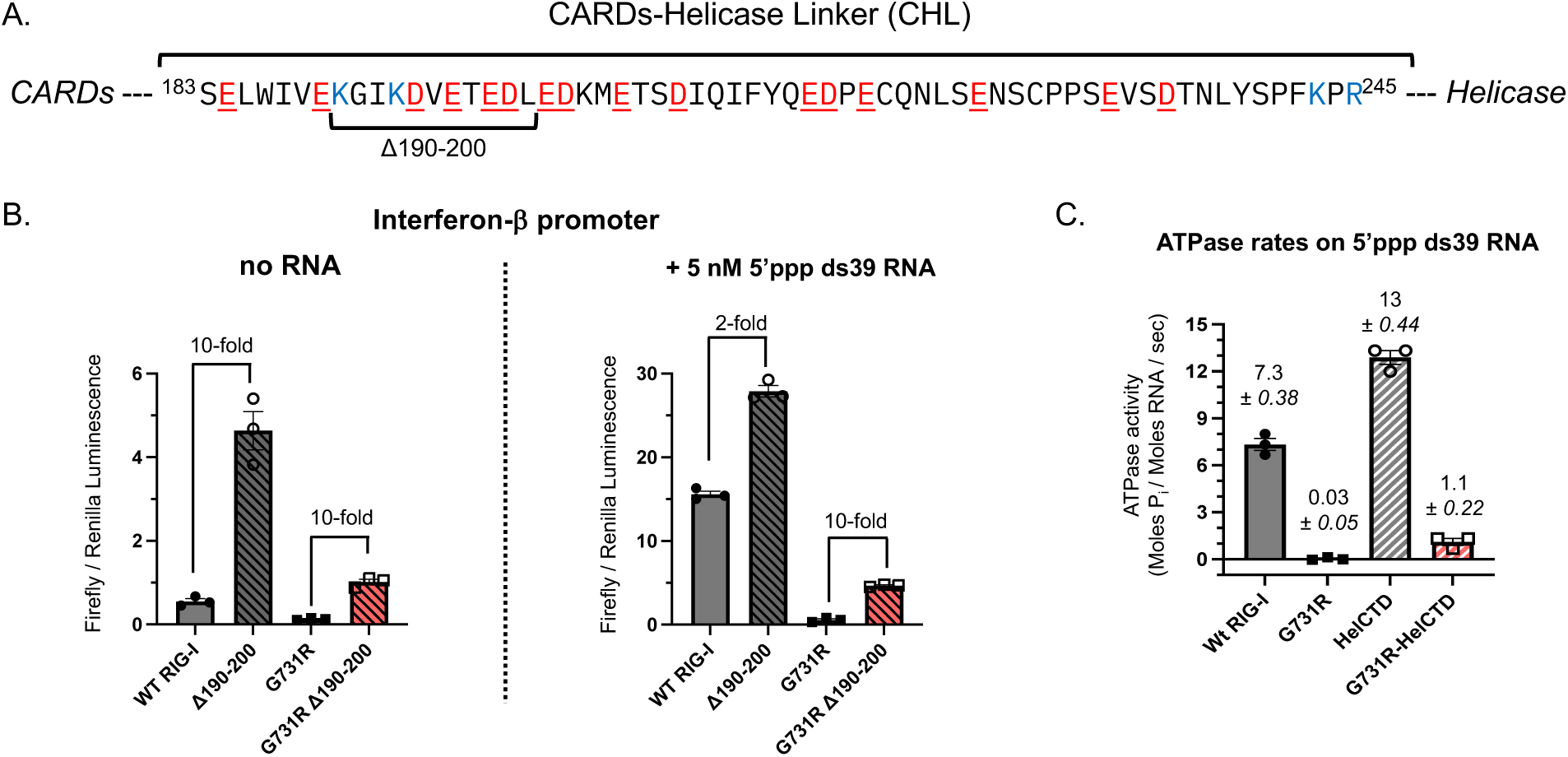
**The CHL regulates IFN-response and ATPase activities of G731R.** (A) Amino acid sequence of the intrinsically disordered CHL of RIG-I. Negatively charged residues are colored red and underlined; positive charges are colored blue. The Δ190-200 segment is indicated. (B) IFN-β reporter activities of WT RIG-I, Δ190-200 WT, G731R, and Δ190-200 G731R RIG-I in the absence and presence of 5 nM 5’ppp ds39 RNA (technical replicates, n=3). The fold-change from the 10 aa deletion in WT RIG-I and G731R is shown. (C) Partial rescue of the ATPase function of G731R upon deletion of the CHL-CARDs domains. The ATPase activities of full-length and HelCTD constructs of WT RIG-I and G731R were measured at 1 µM protein in the presence of 5 nM 5’ppp ds39 RNA. Mean rates of hydrolysis (Moles P_i_ / Moles RNA / sec) are reported (technical replicates, n=3).

Interestingly, deletion of the entire CHL and CARDs regions substantially increased the ATPase function of G731R (Figure 5C). The ATPase rate of full-length G731R in the presence of 5’ppp ds39 was 240-fold slower than WT RIG-I, whereas the rate of G731R HelCTD (Helicase-CTD) was only 12-fold slower than WT HelCTD. Together, these results indicate that the CARDs-CHL module previously shown to stabilize the autoinhibited state of RIG-I^43^ also plays a key role in stabilizing the inactive conformation of G731R, and that disrupting this gating system can partially relieve its loss-of-function phenotype.

### G731R mutation traps RIG-I in a kinetically unstable “CTD-mode”

The studies thus far indicate that G731R adopts a different conformation than WT RIG-I on 5’ppp RNA. Previous biochemical studies have shown that RIG-I forms two distinct 5’ppp RNA-bound conformations, distinguishable by their lifetimes on RNA: “CTD-mode” and “Helicase-mode”^18,43^. The “CTD-mode” is the first intermediate in the RIG-I RNA binding pathway, formed by rapid binding of the CTD to the 5′ppp RNA end (Figure 6A). Subsequently, the helicase domains encircle the RNA to adopt the “Helicase-mode” conformation, leading to CARD exposure. These terms reflect the kinetic behaviors of isolated RIG-I domains, with “CTD-mode” resembling the RNA-bound lifetime of the CTD alone, and “Helicase-mode” resembling the combined Helicase-CTD construct^18^. To determine whether the G731R mutation affects the conformational distribution and kinetic properties of the RIG-I-RNA complexes, we measured the RNA off-rate kinetics using the rapid stopped-flow methods (Figure 6B, STAR Methods).

**Figure 6.**
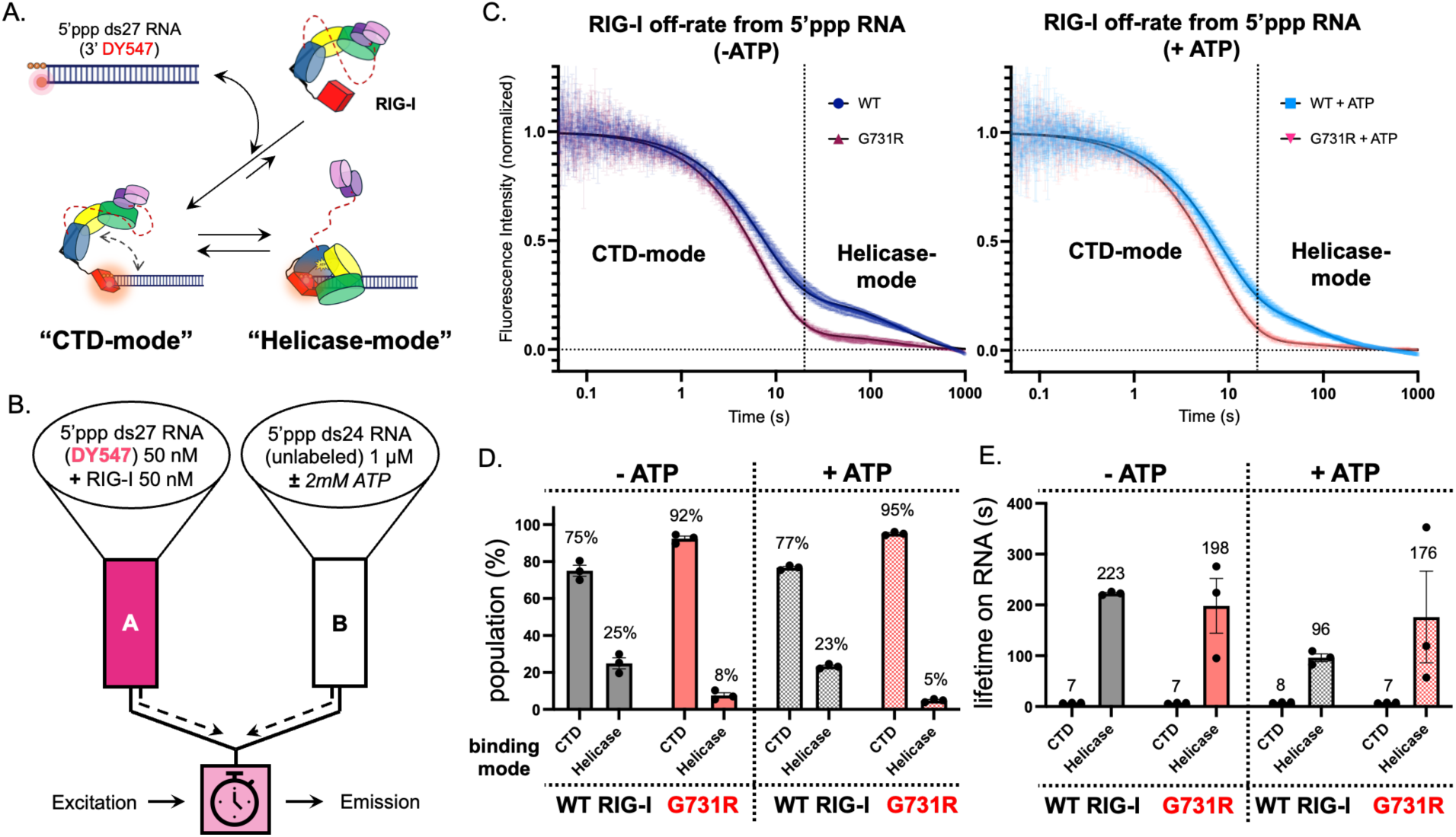
**G731R RIG-I mutant is trapped in the “CTD-mode” RNA-bound conformation.** (A) RIG-I forms kinetically distinguishable “CTD-mode” and “Helicase-mode” conformations on 5’ppp RNA. (B) Experimental setup to measure the off-rate of RIG-I from 5’ppp RNA using a rapid stopped-flow instrument. (C) Biphasic kinetics of WT RIG-I and G731R dissociation from 5’ppp ds27 RNA with and without ATP. The time axis is shown on a log scale to clearly show the two phases. The dissociation kinetics (average of n=3) best fit to the sum of two exponentials (**equation 4**), providing slow and fast off-rates and the corresponding amplitudes plotted in panel D. (D) The “CTD-mode” and “Helicase-mode” phase amplitudes from data in panel C in the absence and presence of ATP are shown as percent population from three biological replicates with SEM. (E) The lifetimes (1/off-rate) in seconds of the “CTD-mode” and “Helicase-mode” phases are shown in the absence and presence of ATP from three biological replicates with SEM.

The off-rate kinetics of WT RIG-I from a DY547-labeled 5’ppp ds27 RNA were biphasic, as expected (Figure 6C), with ∼75% of the molecules dissociating rapidly and 25% dissociating more slowly (Figure 6D). The rapidly dissociating “CTD-mode” conformation had a lifetime of ∼ 7 s, and the “Helicase-mode” had a lifetime of ∼200 s. In contrast to WT RIG-I, only ∼8% of the G731R molecules were found in the “Helicase-mode” (∼198 s), while ∼92% remained in the “CTD-mode”. These data suggest that the G731R-RNA complex is strongly biased toward the “CTD-mode”. Note that the greater variability in the amplitudes and rates of the G731R slower phase is due to the very small proportion of molecules forming the “Helicase-mode”. In fact, because G731R predominantly forms the “CTD-mode” conformation, the off-rate kinetics of the mutant could be fit adequately by a single-exponential model. Addition of ATP altered the dissociation kinetics of WT RIG-I by shortening the “Helicase-mode” lifetime by ∼50%, consistent with ATPase-driven translocation and RNA release, while leaving the “CTD-mode” lifetime unaffected (Figure 6E) (Table S3). By contrast, ATP had no effect on either kinetic phase of G731R, consistent with its lack of ATPase activity.

We conclude that G731R is trapped in the signaling-inactive “CTD-mode” RNA-bound state, unable to efficiently transition to the fully engaged “Helicase-mode” RNA-bound state. This biochemical data and the HDX-MS structural analysis explain the loss-of-function phenotype of G731R. The ability to bind 5’ppp RNA with a high affinity explains the dominant negative phenotype of G731R.

### G731 substitutions lead to both loss-of-function and gain-of-function phenotypes

To investigate whether the loss-of-function phenotype of G731R was specific to the arginine mutation, we mutated G731 to lysine (K), glutamic acid (E), alanine (A), leucine (L), and phenylalanine (F), thereby examining the effects of varying side chain charge and size. IFN-β reporter assays revealed that bulky positively charged (G731K) and negatively charged (G731E) mutations resulted in loss-of-function phenotypes, while G731F showed partial activity (Figure 7A). By contrast, the small, uncharged G731A and G731L sidechains were fully active in the presence of 5’ppp ds39 RNA and, surprisingly, showed higher basal signaling than WT RIG-I, like the gain-of-function SMS mutants of RIG-I^46,47^. Thus, steric hindrance from bulky substitution at G731 may explain the conformational shift to the inactive state. Indeed, modeling using the PyMOL mutagenesis wizard predicted steric clashes between R731 and nearby beta strands in the Hel2 domain (aa 711-715, 737-742) when modeled into the ATPase-competent RIG-I structure (PDB: 4A36) (Figure 7B). Visually, there appears to be an inverse relationship between the degree of predicted steric hindrance (modeled in PyMOL) and IFN-β signaling activity, with bulky charged substitutions showing the greatest clashes and lowest signaling (Figure 7B). Overall, these findings indicate that a bulky and charged residue at position G731 traps RIG-I in a CARDs-protected signaling-inactive “CTD-mode” conformation, preventing its transition to the active “Helicase-mode” state (Figure 7C).

**Figure 7.**
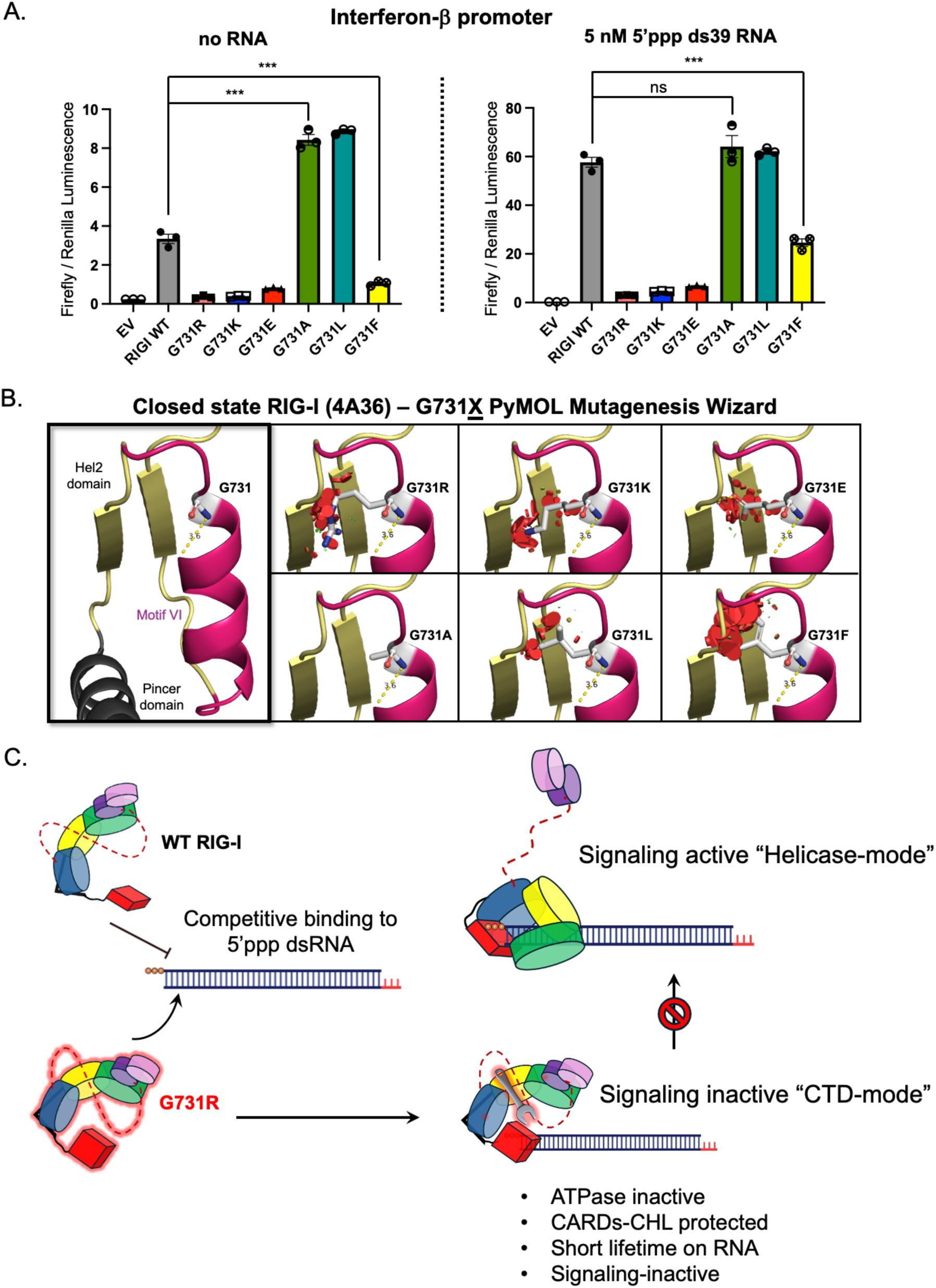
**Bulky and charged substitutions at G731 correlate with loss-of-function.** (A) IFN-β responses of various G731<u>X</u> mutants with and without RNA were measured by dual luciferase reporter assay (technical replicates, n=3). (B) PyMOL Mutagenesis Wizard analysis predicts clashes with bulky substitutions of G731 in the closed state of RIG-I (4A36). The visible red disks indicate pairwise overlap of atomic van der Waals radii. (C) Mechanistic model of G731R mutation locking RIG-I in an inactive CTD-mode RNA-bound conformation, where the CARDs-CHL are not exposed. The high affinity of G731R for the 5’ppp RNA competitively interferes with the WT RIG-I binding to the RNA.

Interestingly, while the loss-of-function G731R RIG-I variant we identified in our patient is novel, an extremely rare single-nucleotide polymorphism for G731E, which we also determined to be loss-of-function, was previously identified in one individual of unknown affected status in the general population database (gnomAD v4.1.0, accessed July 2025). Other potential single-nucleotide polymorphisms, including those that substitute G731 with alanine or valine, have yet to be identified in humans. These variants are predicted to have high CADD scores (>20), but their AlphaMissense scores, which are based upon protein structural predictions^52^, classified G731A (0.308) as likely benign (Table S4) even though it possesses a significant gain-of-function in our assay (Figure 7A). Taken together, these findings not only highlight the critical role of the G731 residue in regulating RIG-I enzymatic activity but also that different substitutions at this position may lead to distinct clinical outcomes, with important diagnostic ramifications.

## DISCUSSION

Here, we identified a heterozygous RIG-I G731R dominant negative variant in a patient with critical COVID-19 pneumonia. We found that the G731R variant is not only a biochemically loss-of-function variant but also strongly interferes with the function of WT RIG-I, leading to severely compromised RIG-I signaling in the heterozygous patient. To date, no patients with complete RIG-I functional deficiency have been reported; however, in mice, RIG-I knockout is embryonically lethal ^29,30^, suggesting that its physiological role extends beyond antiviral signaling. Patients with a milder partial RIG-I deficiency, caused by haploinsufficiency or a common eQTL, were susceptible to severe influenza and acute Hepatitis E virus infections^25–27^. This RIG-I G731R patient, despite being otherwise healthy, developed critical COVID-19 pneumonia, indicating that RIG-I plays a crucial yet not fully understood protective role in response to SARS-CoV-2 in humans.

The mechanistic basis for the loss-of-function phenotype for the RIG-I G731R variant was elucidated using biochemical and biophysical approaches. Our studies showed that G731R binds to 5’ppp RNA with a high affinity, but it fails to activate the ATPase or the IFN-β response activities. The G731 residue is located in the Hel2 domain within helicase motif VI, positioned between two catalytic arginines that are critical for ATP hydrolysis. Substituting G731 with a bulky residue likely interferes with the positioning of the catalytic arginines. The helicase motif VI is present at the interface of Hel1 and Hel2, which undergoes a substantial structural change when RIG-I binds RNA and transitions from the autoinhibited open conformation to the ATPase-active closed conformation (Figure S10A). Existing structures of RIG-I indicate that during this process, motif VI transitions from a loop to an alpha-helix, a necessary process to align the catalytic arginines for ATP hydrolysis (Figure S10B). Mutating G731 to a bulky arginine likely disrupts this folding process, locking it in the initial “CTD-mode and preventing the transition to the “Helicase-mode” closed state necessary for activating the ATPase and signaling functions of RIG-I (Figure 7C).

The lack of ATPase activity cannot be the sole reason for the impairment in the signaling function of the G731R variant. SMS disease variants with mutations in motif I (C268F) and motif II (E373A) are ATPase-inactive 5’ppp RNA binders but show enhanced signaling activity in the absence of viral RNA compared to WT RIG-I^18,22,46^. Previous RNA off-rate kinetics studies of the SMS mutants showed that the inability to hydrolyze ATP results in long-lived “Helicase-mode” complexes on 5’ppp RNA in the presence of ATP^18^. The lack of ATPase-mediated proofreading led to stable complexes of the SMS mutant RIG-I proteins on nonspecific host RNAs, accounting for the unchecked inflammatory symptoms of the disease^46,47^. The G731R variant behaves differently from both WT RIG-I and the SMS mutants. While G731R does not hydrolyze ATP, it fails to form the “Helicase-mode” RNA complex like the SMS mutants ^18^. Instead, it is trapped in the “CTD-mode” conformation on 5’ppp RNA. As shown previously, the “CTD-mode” is an intermediate in the RNA-binding pathway of RIG-I, resulting from high-affinity binding of the CTD domain to the 5’ppp RNA end^18^. Thus, G731R is locked in an intermediate 5’ppp RNA-bound state, unable to transition to the signaling active RNA-bound state (Figure 7C). HDX-MS studies showed that, unlike in WT RIG-I, where RNA binding induces solvent exposure of the CARDs and CHL, these regions remained solvent-protected in G731R. Thus, the CARDs are protected in the “CTD-mode” conformation, which explains the impaired downstream signaling activity of G731R.

The dominant negative phenotype of G731R arises from competition with WT RIG-I for the 5’ppp RNA. The high affinity of G731R for the 5’ppp RNA enables it to compete effectively with WT RIG-I when coexpressed, preventing WT from binding to the viral RNA and initiating a signaling response. Thus, the studies of this loss-of-function mutant critically reveal that 5′ppp RNA binding alone is insufficient for RIG-I CARD activation.

For signaling capability, the helicase domains must engage with the RNA to form a long-lived closed “Helicase-mode” complex, enabling full exposure of the CARDs-CHL for interactions with downstream adapters.

Mutations like G731R, which stabilize transient intermediate states in the RNA binding pathway (Figure 7C), provide valuable insights into these otherwise inaccessible states. Studying these mutations not only elucidates disease mechanisms but also helps pinpoint key regulatory elements that control RIG-I signaling fidelity. Such regulatory elements are potential targets for therapeutic intervention, enabling the design of agonists and antagonists that modulate immune responses. Our observation that the CHL deletion can partially activate G731R suggests that targeting the CHL or ATPase motifs with small molecules or peptides could offer a means to fine-tune RIG-I activity and control immune responses. Furthermore, in heterozygous patients carrying dominant-negative variants like G731R, therapeutic strategies may not necessarily aim to restore function to the mutant itself, but rather to offset its suppressive effects on the WT allele. One potential approach would involve developing RIG-I agonists that mimic the ATPase-deficient SMS variants, promoting WT RIG-I binding and signaling on nonspecific host RNAs. This compensatory mechanism may overcome competitive inhibition by the mutant and restore antiviral signaling capacity.

Our studies showed that point mutations that affect RIG-I conformation and RNA activity can lead to opposing gain-of-function or loss-of-function phenotypes, demonstrating the crucial link between RIG-I conformational dynamics and its functional activity. These studies underscore the importance of complementing computational predictions of pathogenicity with biochemical validation when establishing causality for patient-derived variants. As has been observed for certain *STAT3* mutations^53^, experimentally validating the biochemical impact of different amino acid substitutions, even at the same residue, is required. Despite its high CADD score and observed gain-of-function phenotype (Figure 7A), AlphaMissense incorrectly predicted G731A to be benign (Table S4). This highlights a potential limitation of AlphaMissense in accurately predicting the impact of all variants in specific proteins. These findings suggest that different mutations at this crucial residue can lead to divergent clinical outcomes, ranging from gain-of-function, as seen in SMS, to loss-of-function associated with increased susceptibility to viral infections. Such distinctions not only underscore the need for tailored treatment approaches in genetically variable patients, but also highlight the plasticity of RIG-I modulation through either activation or suppression at regulatory checkpoints within its biochemical activation pathway.

### Limitations of the study

Our conclusions about the *in vivo* pathogenicity of the G731R variant are based on one patient with severe RIG-I antiviral deficiency, with limited availability of patient samples for experimental testing. Therefore, we do not know the full spectrum of disease, including the possibility of variable disease penetrance, which has been observed for other inborn errors of immunity involving genes of the type I IFN pathway^24^. Since the G731R variant is novel, and embryonic lethality is seen in RIG-I knockout mice^29,30^, it is unlikely that additional patients with the same variant will be easily identified for further study. Nevertheless, our patient’s history suggests that in humans, RIG-I is important for antiviral immunity. This is supported by the detailed molecular mechanism we have delineated by which the variant impacts recognition of the RIG-I dsRNA ligand for type I IFN signaling. While our HDX studies allowed us to gain insights into the structural changes induced by the G731R mutation, it does not provide the atomic-level detail needed to understand the RNA interactions in the altered state of G731R that leads to impaired signaling function. High-resolution X-ray crystallography or cryogenic electron microscopy of G731R can potentially offer such insights and inspire structure-based drug discovery efforts.

## RESOURCE AVAILABILITY

### Lead contact

Further information and requests for resources and reagents should be directed to and will be fulfilled by the lead contact, Smita Patel.

### Materials availability

The plasmids used in this study will be made available on request. Patient samples are available through A.L. through a material transfer agreement for human materials from the University of Bari Aldo Moro.

### Data and code availability

- The raw data acquired in this study has been deposited at Mendeley Data (link) and are publicly available as of the date of publication.
- The patient’s whole-genome sequencing dataset was deposited in dbGaP under accession number phs002245.v1.p1, which is a controlled access database for qualified investigators.
- No original code has been generated for this publication.
- Any additional information required to reanalyze the data reported in this paper is available from the lead contact upon request.

## Data Availability

All data produced in the present study are available upon reasonable request to the authors.

## ACKNOWLEDGEMENTS

We thank the patient for participating in this research study, Dr. Andrew Snow for scientific discussions and critical comments on the manuscript, and Mary Magliocco for technical assistance. This work was supported by the National Institute of General Medical Sciences (NIGMS) [GM118086 to S.S.P.], the Intramural Research Programs of the National Institute of Allergy and Infectious Diseases [1ZIAAI001265 to H.C.S.], [1ZIAAI001383 to J.M.], [1ZIAAI001312 to S.M.H.]. The contributions of the NIH authors were made as part of their official duties as NIH federal employees, are in compliance with agency policy requirements, and are considered Works of the United States Government. However, the findings and conclusions presented in this paper are those of the authors and do not necessarily reflect the views of the NIH or the U.S. Department of Health and Human Services.

## AUTHOR CONTRIBUTIONS

Conceptualization, M.S., H.J., W.T., P.R.G., J.M., H.C.S., S.S.P.; Methodology, M.S., H.J., E.G., S.J.N., P.G., H.C.S., S.S.P.; Formal Analysis, Y.Z.; Investigation, M.S., T.M., H.J., E.G., S.J.N., B.D.P., W.T., P.H., C.A., A.S., P.L., E.R.S., L.B.R. ; Resources, A.S., P.L., A.L.; Writing, - Original Draft, M.S., H.J., H.C.S., S.S.P.; Writing – Review and Editing, M.S., H.J., W.T., P.H, J.M., H.C.S., S.S.P.; Visualization, M.S., H.J., Y.Z.; Data Curation, M.S., B.D.P., Y.Z.; Supervision, S.M.H., C.L.D., H.C.S., S.S.P.; Project Administration, A.L., H.C.S.; Funding Acquisition, J.M., H.C.S., S.S.P.

## STAR METHODS

### Patient information and sequencing

The patient provided written informed consent for his enrollment and participation in research protocols 1563 (Bari, Italy) and 06-I-0015 (NCT00246857; NIH, USA), which were approved by their respective local institutional review boards (IRB). He was classified as critically ill according to the Diagnosis and Treatment Protocol for Novel Coronavirus Pneumonia based upon the following criteria^55^: mechanical ventilation (CPAP, BiPAP, intubation, hi-flow oxygen); or septic shock; or organ damage requiring admission in the ICU; or severe patients with oxygen saturation at rest 93% or lower OR respiratory rate >30/min OR PaO2/FiO2 <300. Genomic DNA was extracted from frozen whole blood from the patient. Whole genome sequencing was performed using Illumina NovaSeq 6000 sequencing system (Illumina). All sequencing reads were aligned with hg19 genome assembly with Burrows–Wheeler Aligner (BWA) and the variant calling were done by the Genome Analysis Toolkit (GATK) best-practice pipeline.

To confirm the *RIG-I* mutation in the patient, genomic DNA was isolated from the patient’s blood leukocytes and amplified by PCR using forward primer 5′-ACCAGCATTACTAGTCAGAAGGA −3′ and reverse primer 5′-ACAGCTCAACTTTCCTGGGA −3′, followed by Sanger dideoxy sequencing from both directions.

### Autoantibodies testing

Serum samples from the patient were screened for autoantibodies against IFN-α, IFN-β, and IFN-ω using a multiplex particle-based assay, as previously described^56^. Using this methodology, the cutoff values (mean ± 3 standard deviations) for healthy donors serving as negative controls (n=1099) are fluorescence intensities on the Luminex machine of <1,310, <386, and <1,387 for anti-IFN-α, anti-IFN-β, and anti-IFN-ω autoantibodies, respectively.

### Plasmids and mutagenesis

Human *RIG-I* cDNA corresponding to NCBI reference sequence NM_014314.3 was purchased from SinoBiological (clone HG17547-U) and subcloned into pcDNA3.1 (ThermoFisher). The G731R mutant was generated by site-directed mutagenesis through PCR amplification of full-length plasmids encoding WT RIG-I with a primer pair containing the corresponding mutation as previously described^57^.

### Interferon-β Reporter cell signaling assays

HEK293T RIG-I^-/-^ cells were grown at 5% CO_2_ and 37°C in 2 mL of DMEM with 10% FBS to 70% confluency and co-transfected with 500 ng firefly luciferase reporter plasmid (pLuc125), 100 ng renilla luciferase plasmid (pRL-TK), and 50 ng of a plasmid carrying either the WT RIG-I gene or mutant construct under the constitutively active CMV promoter (pcDNA 3.1) or EF1-⍺ promoter (pEF-BOS), as indicated in the text. The firefly luciferase gene is under the control of the interferon-β promoter, and the renilla luciferase plasmid is under the control of the constitutively active TK promoter. The plasmid transfections were performed with X-tremeGENE HP DNA Transfection Reagent (Roche). Cells were replated in 96-well plates the next day at 250,000 cells/mL density and transfected with 0, 5, or 50 nM 5’ppp ds39 RNA using Lyovec transfection reagent (InvivoGen). After 20 hours, the activities of firefly and renilla luciferases were measured sequentially with the Dual-Luciferase Reporter Assay System (Promega).

Data was collected in triplicate sets, and a ratio of firefly to renilla luciferase activity were calculated to indicate relative interferon expression levels. Competition assay was performed by titrating CMV-G731R plasmid against 50 ng EF1-⍺ WT RIG-I in the presence or absence of 5 nM 5’ppp ds39 RNA. Total plasmid concentration was kept constant by substitution with empty vector. The ratio of firefly to renilla (Y) were plotted against G731R plasmid (X) concentration. The inverse hyperbolic dependence was fit to equation 1 to obtain the half maximal inhibitory concentration (IC_50_). Top represents the Y-intercept in the absence of G731R inhibition, and Bottom represents the baseline of signaling at maximum inhibition.

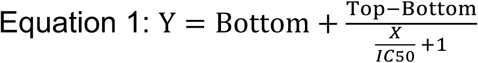

In some experiments, 293T cells were seeded at 100,000 cells per well in 48-well plates one day prior to transfection. Cells were transiently transfected using Lipofectamine 2000 (Invitrogen) with firefly luciferase reporter plasmid driven by human IFN-β promoter (100 ng) and a constitutively expressed Renilla luciferase reporter plasmid (10 ng) together with WT or G731R RIG-I constructs (20 ng). Six hours after transfection, cells were infected with hPIV (MOI 3) and cultured for an additional 20 hours before cell lysis for Luciferase activity. Dual luciferase activities were measured per the manufacturer’s protocol (Promega) on a microplate reader (BMG Labtech Fluostar Omega). Firefly luciferase activity for G731R was normalized to Renilla Luciferase activity and then reported as fold change relative to WT. To test for dominant negative effect of G731R on WT RIG-I, 293T cells were co-transfected with either EV or G731R constructs (5 to 20 ng), along with RIG-I WT (5 ng) for 6 hours. Luciferase activity was induced by infecting with hPIV (MOI 3) before lysing cells 20 hours later. Firefly luciferase activity was first normalized to Renilla luciferase activity and then shown as fold change relative to EV.

### Western Blots

Total protein lysate samples were prepared by pelleting and lysing 250,000 cells in 40 μL of 2× SDS sample buffer (62.5 mM Tris, pH 6.8, 1% SDS, 15% glycerol, 2% β-mercaptoethanol, and 0.005% bromophenol blue) and then boiled for 10 min. Proteins were separated by SDS PAGE and transferred onto 0.2 μm PVDF membrane. The membrane was blocked with 5% nonfat milk in 1X PBST and immunoblotted with 1:500 anti-RIG-I (CST D14G6 Rabbit mAb #3743S) and 1:3000 anti-actin (CST 8H10D10 Mouse mAb #3700S) antibodies at 4°C overnight. After washing, the membrane was immunoblotted with HRP-conjugated secondary antibodies (1:10,000, goat anti-rabbit (RIG-I) or goat anti-mouse (β-actin) IgG) for 1 hour. Blots were developed by using ECL western blotting substrate (ThermoFisher, Pierce) on a BioRad ChemiDoc system. In some experiments, protein lysates from 293T cells transfected with equal amounts of RIG-I WT or G731R plasmids were separated on 3–8% Tris-Acetate gels and then transferred to nitrocellulose membranes using the TransBlot Turbo system (Bio-Rad).

Membranes were blocked with 5% non-fat dry milk in PBST, then incubated overnight with anti-RIG-I antibody (1:1000; Proteintech, 20566-1-AP). After washing, membranes were incubated with HRP-conjugated goat anti-rabbit secondary antibody (1:10,000; Jackson ImmunoResearch), and signals were detected using ECL Western blotting substrate. Membranes were subsequently re-probed with anti-β-tubulin antibody (1:1000; Cell Signaling Technology, #2146), followed by the appropriate HRP-conjugated secondary antibody for signal detection.

### Protein expression and purification

RIG-I constructs were cloned in the protein expression vector pET28 SUMO and expressed as SUMO fusion proteins in *Escherichia coli* strain Rosetta (DE3) (Novagen). The soluble fraction was purified through a HisTrap HP column (GE Healthcare), followed by Ulp1 protease digestion to remove the 6x His-SUMO tag. The filtered soluble fraction was further purified through a heparin Sepharose column (GE Healthcare) to remove any contaminating nucleic acid. The purified protein was dialyzed into 50 mM HEPES pH 7.5, 50 mM NaCl, 5 mM MgCl_2_, 5 mM DTT, and 10% glycerol overnight at 4°C, and finally passed through a Superdex 200 10/300 Increase size exclusion column (GE Healthcare) before being snap frozen in liquid nitrogen and stored at −80°C.

### RNA synthesis by *in vitro* transcription

5’ triphosphate 39-mer ssRNA was transcribed *in vitro* using a bacteriophage T7 RNA polymerase system^42^. 400 µl transcription reactions were run for 3 h at 30°C after mixing the required reaction components, including 250 mM HEPES pH 7.5 buffer, 30 mM MgCl2, 40 mM DTT, 2 mM spermidine, 5 µg inorganic pyrophosphatase, 50 U RNasin, 2 µM dsDNA promoter and template sequence, 5 mM ATP, GTP, and UTP, and 1 µM T7 RNA polymerase, purified as previously described^58^. Reactions were quenched with 75 mM EDTA. The 39-mer ssRNA products were purified by gel electrophoresis on 1.5 mm × 30 cm 40% acrylamide and 6 M urea gel in 1.5× TAE. Electrophoresis was conducted at 500 V for 4 h, and the resulting ssRNA was identified by UV shadowing, the band was excised, and RNA was extracted from the gel using a Whatman Elutrap electroelution system in 1.5x TAE buffer run at 150 V for 6 h. The resulting ssRNA was concentrated with ethanol precipitation and stored at –80°C. The 39-mer ssRNA was analyzed for purity using LC–MS (Novatia, LLC; Newton, PA).

The 5’OH complementary fluorescent, or non-fluorescent, ssRNA was chemically synthesized (Dharmacon). Fluorescein-labeled 5’ppp 10bp HP RNA was chemically synthesized (TriLink).

### Electrophoretic Mobility Shift Assay (EMSA)

EMSAs were performed by incubating 0, 20, 40, and 80 nM WT RIG-I or G731R variant with 20 nM 5’ppp ds39 RNA in ATPase buffer (50 mM MOPS pH 7.4, 5 mM DTT, 5 mM MgCl_2_, 0.01% Tween20) for 30 minutes at 4°C. 2 mM ATP was optionally added to each reaction 15 min before loading the sample into the gel. 3 µl loading buffer (15% Ficoll 400 in Tris-borate buffer, pH 8.0) was added to the samples prior to loading on a 4–16% gradient Native PAGE gel (Invitrogen) and run at 4°C. Gels were scanned at 532 nm using a Typhoon FLA 9500 laser-based scanner (GE).

### Fluorescence-based intensity and polarization RNA-binding assays

Fluorescence intensity and polarization measurements were performed on a Tecan Spark microplate reader in a 384-well black plate at 25°C. A monochromator set the excitation wavelength at 485 nM and the emission wavelength at 535 nm, with a 20 nm bandwidth. To measure 5’ppp 10bp HP RNA binding affinity, protein was serially titrated in ATPase buffer (50 mM MOPS pH 7.4, 5 mM DTT, 5 mM MgCl_2_, 0.01% Tween-20) and incubated with a constant 2 nM fluorescein-labeled 5’ppp 10bp HP RNA for 15 min at 25°C. Readings were taken in the presence of 500 µM ATP. To obtain K_D_ values, fluorescence intensity and miliPolarization values from triplicate data sets were plotted as a function of protein concentration. Fluorescence intensity or polarization (Y) were plotted against protein (X) concentration, and the hyperbolic dependence was fit to equation 2 to obtain the binding amplitudes (Bmax) and K_D_ (Kd). C is the intercept representing the fluorescence intensity of the RNA in the absence of protein.

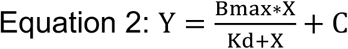

5’ppp ds39 RNA competition assay was performed by titrating unlabeled 5’ppp ds39 RNA against 10 nM equimolar RIG-I-bound 5’ppp 10bp HP RNA (FAM). Changes in fluorescence polarization of the 5’ppp 10bp HP RNA (FAM) were monitored (Y) and plotted against 5’ppp ds39 RNA (X) concentration. The inverse hyperbolic dependence was fit to equation 1 to obtain the half maximal inhibitory concentration (IC_50_). From these experiments, the Cheng-Prusoff equation (equation 3) was used to obtain 5’ppp ds39 RNA K_i_ values, which are representative of RIG-I’s binding affinity for 5’ppp ds39 RNA (K_D_).

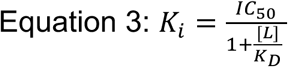

### Radiometric ATPase Assay

ATP hydrolysis was measured at variable 5’ppp ds39 RNA and RIG-I concentrations in the presence of 2 mM ATP (spiked with [γ-^32^P] ATP). The ATPase reaction was measured after 0 (prequench), 20 and 40 minutes of reaction time in ATPase buffer (50 mM MOPS pH 7.4, 5 mM DTT, 5 mM MgCl_2_, 0.01% Tween-20) at 25°C. Reactions were stopped at each time point using 4 N formic acid and analyzed by PEI-Cellulose-F TLC (Merck) developed in 0.4 M potassium phosphate buffer (pH 3.4). TLC plates were exposed to a phosphoimager plate, imaged on a Typhoon phosphor-imager, and quantified using the ImageQuant software. The molar [Pi] generated during the reaction time intervals was plotted against time and fit to a linear equation to obtain the ATPase rate (Moles Pi/s).

ATPase-based RNA competition assay was performed by titrating G731R protein against 50 nM WT RIG-I in the presence of 20 nM 5’ppp ds39 RNA. ATPase rates were divided by the 20 nM RNA (Y) and plotted against G731R protein (X) concentration. The inverse hyperbolic dependence was fit to equation 1 to obtain the half maximal inhibitory concentration (IC50). Top represents the Y-intercept in the absence of G731R inhibition, and Bottom represents the baseline of ATPase at maximum inhibition.

### Mant-ATP binding assay

Fluorescence intensity measurements were performed on a Tecan Spark microplate reader in a 384-well black plate at 25 ^◦^C. A monochromator set the excitation wavelength at 290 nM and the emission wavelength at 425 nm, with a 20 nm bandwidth. Mant-ATP (Invitrogen Life Technologies) was serially titrated in ATPase buffer (50 mM MOPS pH 7.4, 5 mM DTT, 5 mM MgCl_2_, 0.01% Tween-20), and baseline fluorescence readings were taken. 500 nM protein was then added to each well containing Mant-ATP and incubated for 15 min at 25°C before a second fluorescence measurement was taken. The difference in fluorescence intensity readings before and after adding protein was plotted as a function of Mant-ATP concentration. The Δfluorescence intensity (Y) was plotted against Mant-ATP (X) concentration, and the hyperbolic dependence was fit to equation 2 to obtain the binding amplitudes (Bmax) and K_D_ (Kd). C is the intercept representing the fluorescence intensity of the protein alone.

### Hydrogen-Deuterium Exchange – Mass Spectroscopy (HDX-MS)

*HDX-MS analysis:* Ten micromoles of WT RIG-I or G731R mutant in buffer (50 mM HEPES, pH 7.4, 150 mM NaCl, 5% glycerol, 5 mM MgCl_2_, 2 mM DTT) were optionally incubated with 5’ppp 10bp HP RNA ligand at a 1:1.2 molar ratio (protein: ligand) for 1 h before the HDX reactions at 4 °C. Five microliters of protein with or without RNA was diluted into 20 µL D_2_O in exchange buffer (50 mM HEPES, pH 7.4, 150 mM NaCl, 5 mM MgCl_2_, 2 mM DTT) and incubated for various HDX time points (e.g., 0, 30, 60, 300, 600, 900, 1800, and 3600 s) at 4°C and quenched by mixing with 25 µL of ice-cold 4 M guanidine hydrochloride and 1% trifluoroacetic acid. Dmax samples were incubated in D_2_O in the exchange buffer containing 3 M guanidine hydrochloride (50 mM HEPES, pH 7.4, 150 mM NaCl, 5 mM MgCl_2_, 2 mM DTT, and 3 M guanidine hydrochloride) overnight at room temperature. The sample tubes were immediately placed on dry ice after quenching until the samples were injected into the HDX platform. Upon injection, samples were passed through an immobilized pepsin column (2 mm × 2 cm) at 200 µL/min, and the digested peptides were captured on a 2 mm × 1 cm C8 trap column (Agilent) and desalted. Peptides were separated across a 2.1 mm × 5 cm C18 column (1.9 μm Hypersil Gold, ThermoFisher) with a linear gradient of 4–40% CH_3_CN and 0.3% formic acid, over 5 min. Sample handling, protein digestion, and peptide separation were conducted at 4°C. Mass spectrometric data were acquired using an Orbitrap mass spectrometer (Q Exactive, ThermoFisher) with a measured resolving power of 65,000 at m/z 400.

HDX analyses were performed duplicate or triplicate, with single preparations of each protein ligand complex. The intensity weighted mean m/z centroid value of each peptide envelope was calculated and subsequently converted into a percentage of deuterium incorporation. In the absence of a fully deuterated control, corrections for back exchange were made by an estimated 70% deuterium recovery, and accounting for the known 80% deuterium content of the deuterium exchange buffer. When comparing the two samples, the perturbation %D is determined by calculating the difference between the two samples. HDX Workbench colors each peptide according to the smooth color gradient HDX perturbation key (%D) shown in each indicated figure. Differences in %D between −5 to 5% are considered nonsignificant and are colored gray according to the HDX perturbation key. In addition, unpaired t tests were calculated to detect statistically significant (p < 0.05) differences between samples at each time point. At least one time point with a p value <0.05 was present for each peptide in the data set further confirming that the difference was significant.

*Peptide identification:* Peptides were identified using tandem MS (MS/MS) with an Orbitrap mass spectrometer (Q Exactive, ThermoFisher). Product ion spectra were acquired in data-dependent mode with the top five most abundant ions selected for the production analysis per scan event. The MS/MS data files were submitted to Mascot (Matrix Science) for peptide identification. Peptides included in the HDX analysis peptide set had a MASCOT score >20, and the MS/MS spectra were verified by manual inspection. The MASCOT search was repeated against a decoy (reverse) sequence and ambiguous identifications were ruled out and not included in the HDX peptide set. *Data rendering:* The HDX data from all overlapping peptides were consolidated to individual amino acid values using a residue averaging approach. Briefly, for each residue, the deuterium incorporation values and peptide lengths from all overlapping peptides were assembled. A weighting function was applied in which shorter peptides were weighted more heavily, and longer peptides were weighted less. Each of the weighted deuterium incorporation values was then averaged to produce a single value for each amino acid. The initial two residues of each peptide, as well as prolines, were omitted from the calculations.

### Stopped-Flow Off Rate Measurements

RNA off-rates were measured at 25°C using a stopped-flow instrument (Auto-SF 120, Kintek Corp, Austin, Tx). To initiate the reaction, 50 nM 5’ppp ds27 DY547-labelled RNA and 50 nM RIG-I or mutant protein in ATPase buffer (50 mM MOPS pH 7.4, 5 mM DTT, 5 mM MgCl_2_, 0.01% Tween-20) (pre-incubated at 25°C for 10 min) from syringe A was mixed with a 20-fold excess of an RNA trap (unlabeled 5’ppp ds24 palindromic RNA) from syringe B, with or without 2 mM ATP. The fluorescence emission of DY547 was measured using a 570 nm band-pass filter after excitation at 547 nm. The change in fluorescence intensity was plotted as a function of time, and the data were fit to the sum of two exponentials (equations 4) to estimate the off-rates for each unbinding phase.

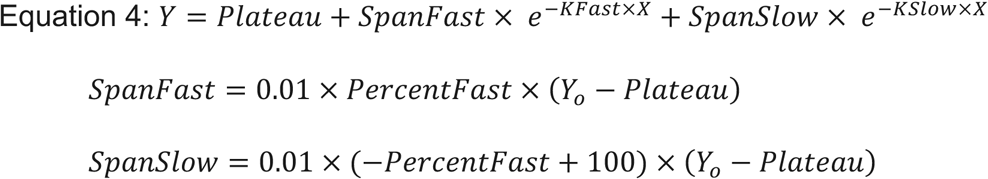

## QUANTIFICATION AND STATISTICAL ANALYSIS

All the experiments were performed as duplicates or triplicates or independent experiments as indicated in the figure legends. Statistical analysis of the difference in data was performed using an unpaired parametric t test design or one-sample t-test in Prism GraphPad. Annotations for statistical differences are indicated in the figures as follows: ns → no significant difference, (*) → p < 0.05, (**) → p < 0.01, (***) → p < 0.001, (****) → p < 0.0001.

