## Supplemental Figures for "Human RIG-I Antiviral Deficiency Caused by a Dominant-Negative Variant Locked in a Signaling-Inactive State"

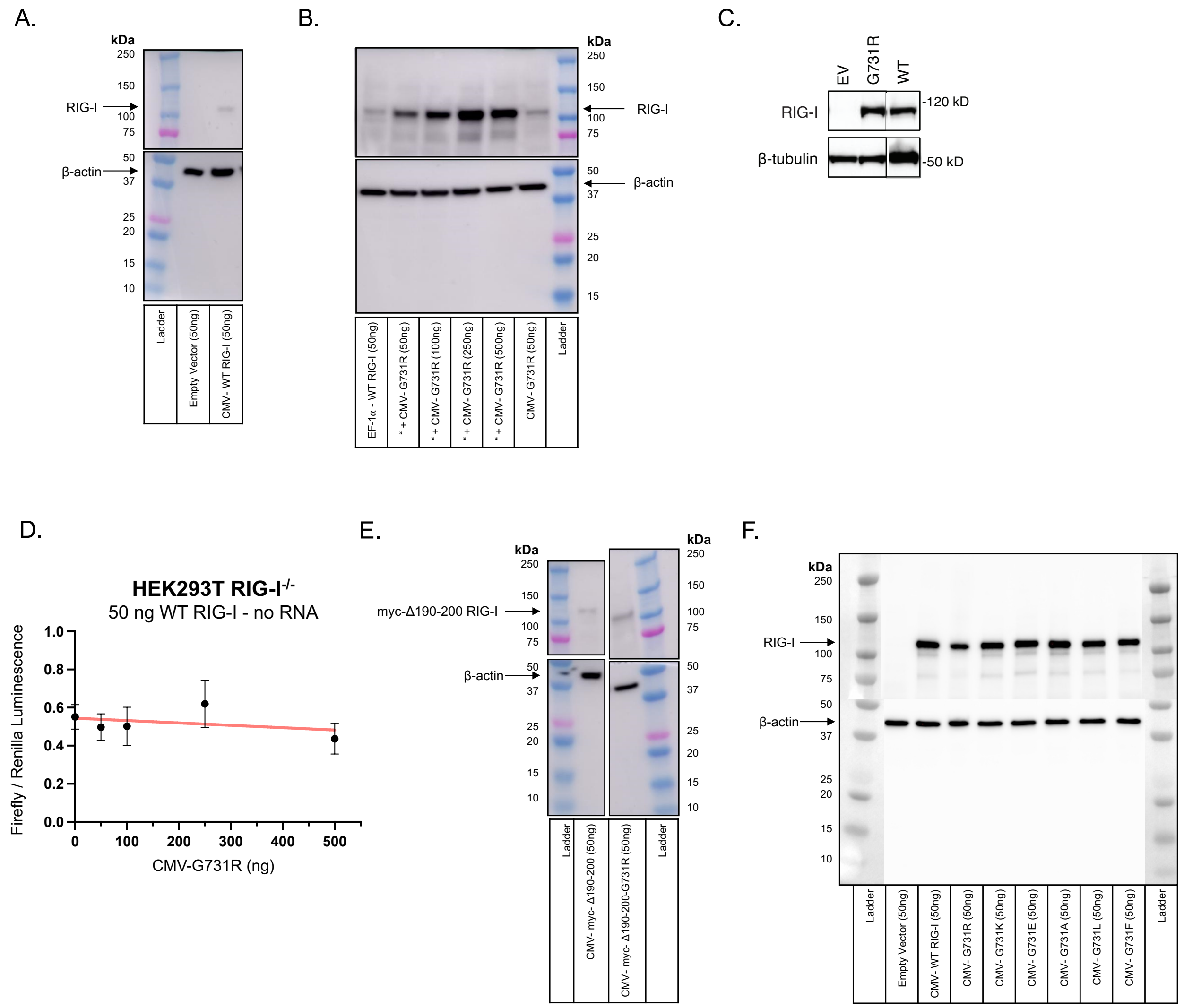


**Figure S1. IFN-β reporter assay western blots and control**

Western blot corresponding to IFN-β reporter assay in HEK293T RIG-I$^{-/-}$ cells for transient expression of (A) CMV- WT RIG-I, (B) EF1-α- WT RIG-I and competing CMV-G731R, (E) CMV- myc-Δ190-200 constructs, and (F) CMV- G731X mutants relative to β-actin. Panels (A) and (B) are related to data in figure 2, (E) is related to figure 6, and (F) is related to figure 7.

(C) Immunoblot assessing expression of RIG-I WT and G731R, corresponding to Figure 2B. Cell lysates from transfections with empty vector (EV), RIG-I WT, or RIG-I G731R were resolved on the same gel. A section of the gel between the WT and G731R lanes was removed, as indicated by the black line.

(D) IFN-β response of constitutively expressed WT RIG-I under EF1-α promoter measured by dual luciferase reporter assay in the absence of RNA, while titrating G731R plasmid, related to data in figure 5. Each condition was performed three times (technical replicates, n=3). The plot shown represents the mean values at each protein concentration with error bars representing the standard error of the mean (SEM), and the curve represents hyperbolic decay fitting of the mean data.

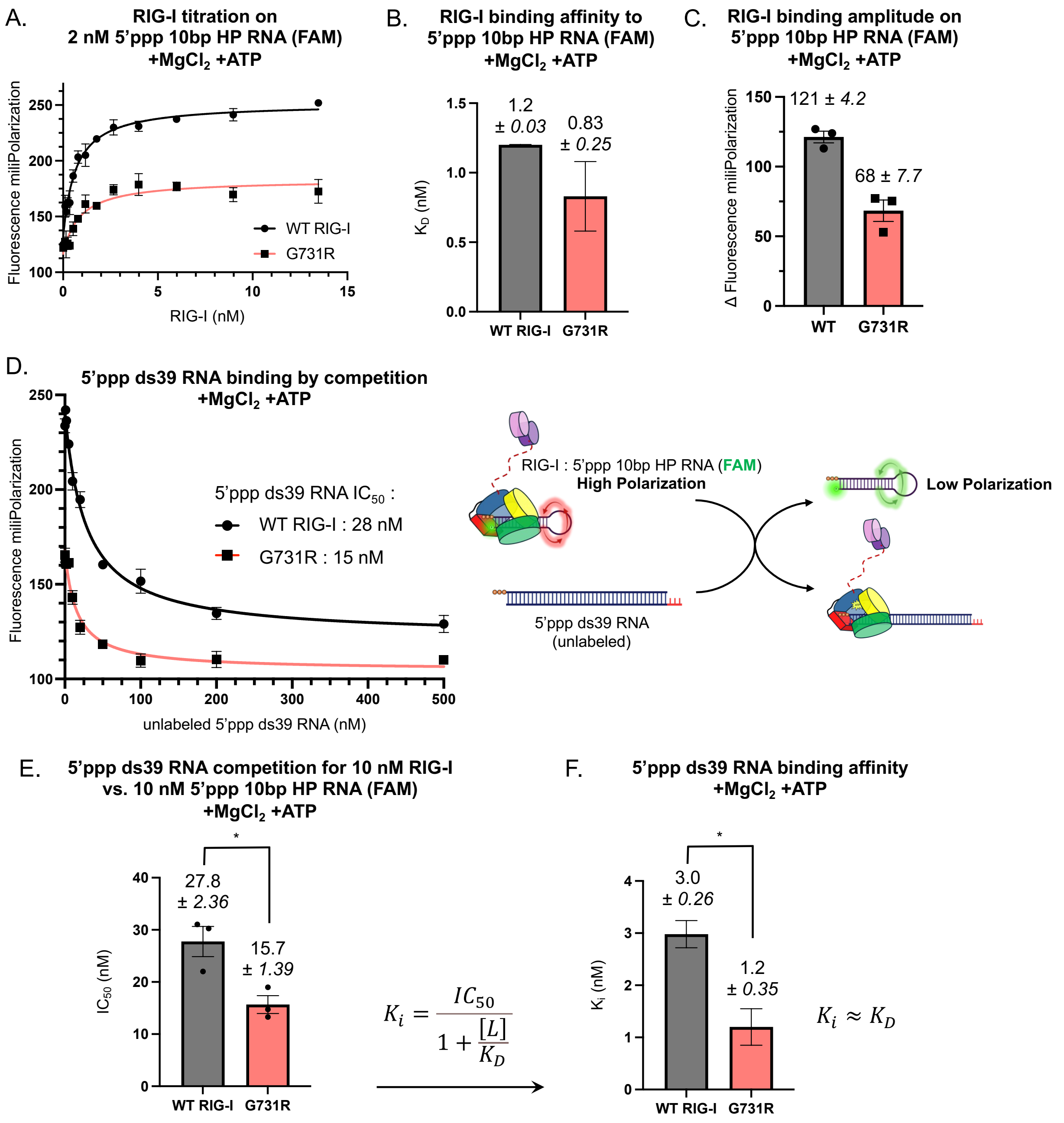


$$K_i = \frac{IC_{50}}{1 + \frac{[L]}{K_D}}$$

$$K_i \approx K_D$$

**Figure S2. Fluorescence-based RNA binding measurements related to RNA binding data in figure 3B.**

(A) Fluorescence polarization (FmP) of fluorescein-labeled 5'ppp 10bp HP RNA in the presence of increasing protein concentration (technical replicates, n=3). The RNA binding data were fit to equation 2 to obtain the FmP amplitude and the RNA $K_D$ values .

(B) 5’ppp 10bp HP RNA binding affinity ($K_D$) measured by changes in fluorescence polarization while titrating RIG-I against constant 2 nM RNA. Error bars represent the standard error of the mean (technical replicates, n=3).

(C) The FmP amplitude values from experiments in panel A are compared (technical replicates, n=3).

(D) 5’ppp ds39 RNA binding to WT or G731R was measured by competition assay. Fluorescence polarization of FAM-labeled 5’ppp 10bp HP RNA (10 nM) bound to RIG-I or G731R (10 nM) was measured while titrating in increasing unlabeled 5’ppp ds39 RNA. The data were fit to equation 1 to obtain the half maximal inhibitory concentration $IC_{50}$. Error bars represent the standard error of the mean (technical replicates, n=3).
(E) 5’ppp ds39 RNA competition ($IC_{50}$) for RIG-I displacement from 5’ppp 10bp HP RNA as monitored by changes in fluorescence polarization while titrating 5’ppp ds39 RNA against 10 nM equimolar RIG-I-bound 5’ppp 10bp HP RNA (FAM). Error bars represent the standard error of the mean (technical replicates, n=3),

(F) Cheng-Prusoff equation is used to obtain 5’ppp ds39 RNA $K_i$ values, which are representative of RIG-I’s binding affinity for 5’ppp ds39 RNA ($K_D$). Error bars represent the propagated standard error of the means. Statistical analysis was performed using unpaired t-test. *, $P<0.05$.

Consolidated HDX differential peptide coverage

| | | | Δ HDX | | | | |
|---|---|---|---|---|---|---|---|
| **Domain** | **Feature** | **Peptide Sequence** | Start Residue | End Residue | wt RIG-I ± 5'ppp 10bp HP RNA | apo G731R vs. WT RIG-I | G731R ± 5'ppp 10bp HP RNA |
| | | FQDYIRKTLDPTYIL | 11 | 25 | 15 (4) | 2 (3)* | -17 (2) |
| | | FQDYIRKTLDPTYILS | 11 | 26 | 15 (4) | -1 (2)* | -21 (3) |
| | | YIRKTLDPTYIL | 14 | 25 | 15 (5) | 2 (2)* | -20 (3) |
| | | SYMAPWFREEEVQ | 26 | 38 | 17 (4) | 1 (2)* | -30 (3) |
| CARD1 (1-91) | | YIQAEKNNKGPMEAATL | 39 | 55 | 11 (4) | -6 (3) | -28 (3) |
| | | LKFLLE | 57 | 62 | | 0 (1)* | 2 (2)* |
| | | QEEGWFRGFLDA | 64 | 75 | 13 (5) | -8 (2) | -17 (3) |
| | | DALDHAGYSGL | 74 | 84 | 8 (2) | -6 (3) | -22 (4) |
| | | DALDHAGYSGL | 74 | 84 | 12 (3) | -6 (3) | -21 (4) |
| | | YEAIESWD | 85 | 92 | 16 (5) | 2 (3)* | -28 (4) |
| | | FKKIEKLEE | 93 | 101 | -4 (7)* | -5 (3) | -40 (3) |
| | CARD2:Hel2i latch (100-115) | YRLLL | 102 | 106 | 11 (3) | 6 (3) | |
| | | KRLQPEF | 107 | 113 | 6 (4) | 6 (2) | -27 (3) |
| | | KTRIIPTDIISDL | 114 | 126 | 17 (4) | 1 (3)* | -33 (3) |
| | | KTRIIPTDIISDL | 114 | 126 | 17 (3) | -2 (3)* | -38 (2) |
| CARD2 (92-185) | | LINQECEE | 130 | 137 | | -2 (3)* | -31 (5) |
| | CARD2:Hel2i latch (145-150) | ILQICSTKGMMAGAEKLVE | 138 | 156 | 22 (3) | 2 (2)* | -19 (4) |
| | | ILQICSTKGMMAGAEKLVE | 138 | 156 | 16 (4) | 2 (1)* | -14 (3) |
| | | LRSDKENWPKTLKLAL | 159 | 174 | 22 (8) | 3 (2)* | -30 (2) |
| | | LRSDKENWPKTLKLAL | 159 | 174 | | 3 (2)* | -29 (4) |
| | | ALEKERNKFSEL | 173 | 184 | 6 (4) | 1 (3)* | -37 (5) |
| | | WIVEKGIKDVETEDL | 185 | 199 | 2 (4)* | -8 (2) | -57 (5) |
| | Electrostatic gate (190-220) | WIVEKGIKDVETEDLEDKMETSD | 185 | 207 | 1 (2)* | | -46 (6) |
| | | EDKMETSDIQIF | 200 | 211 | 4 (6)* | | -25 (3) |
| ID Linker (185-244) | | YQEDPECQNL | 212 | 221 | 3 (4)* | -8 (3) | -54 (7) |
| | | SENSCPPSEVSDTNL | 222 | 236 | 4 (4)* | -7 (2) | -80 (4) |
| | | VSDTNL | 231 | 236 | 2 (3)* | -15 (3) | -88 (5) |
| | | YSPFKPRN | 237 | 244 | 0 (9)* | -12 (2) | -88 (4) |
| | | YSPFKPRNYQLE | 237 | 248 | 0 (6)* | | -55 (4) |
| | | LALPAMKGKNTIICAPTGCGKTFVSL | 249 | 274 | -1 (3)* | -3 (2)* | -33 (2) |
| | | TIICAPTGCGKTF | 259 | 271 | 2 (5)* | -5 (1)* | -56 (4) |
| | RNA binding motifs: | LICEHHLKKFPQGQKGKVVF | 275 | 294 | | -1 (1)* | -23 (2) |
| | | FANQIPVYE | 295 | 303 | -16 (4) | -11 (3) | |
| | motif Ia (300-330) | FERHGYRVTGISGATAENVPVEQ | 313 | 335 | -3 (6)* | -4 (2)* | -54 (4) |
| | | FERHGYRVTGISGATAENVPVEQ | 313 | 335 | -2 (7)* | -5 (1)* | -37 (2) |
| | | IVENND | 336 | 341 | -2 (2)* | 1 (2)* | -7 (3) |
| | | IIILTPQIL | 342 | 350 | -9 (4) | 3 (2)* | -7 (3) |
| Hel1 (244-455) | motif Ic (340-370) | IIILTPQILVNNLKKGTIPSL | 342 | 362 | -12 (5) | -1 (2)* | -36 (3) |
| | | VNNLKKGTIPSLSIF | 351 | 365 | -7 (4) | -2 (2)* | -36 (3) |
| | | MIFDECHNTSKQHPYNM | 368 | 384 | -9 (4) | -1 (2)* | -22 (2) |
| | motif IIa (370-390) | IFDECHNTSKQHPYNMIM | 369 | 386 | -7 (3) | 2 (1)* | -20 (2) |
| | | DECHNTSKQHPYNMIM | 371 | 386 | -9 (4) | 0 (1)* | -27 (3) |
| | | FNYLDQKLGGSSGPLPQVIGL | 387 | 407 | 0 (2)* | 2 (1)* | -22 (2) |
| | | FNYLDQKLGGSSGPLPQVIGL | 387 | 407 | 0 (2)* | 2 (1)* | -20 (2) |
| | | TASVGVGDAKNTDEAL | 408 | 423 | -4 (3)* | -2 (3)* | |
| | | DYICKL | 424 | 429 | | 3 (2)* | 2 (5)* |
| | | DASVIATVKHNLEEL | 434 | 448 | 0 (3)* | 0 (3)* | -8 (2) |
| | | SVIATVKHNLEEL | 436 | 448 | -1 (4)* | 1 (3)* | -12 (2) |
| | Insertion domain linker | LEQVVYKPQKF | 448 | 458 | -4 (5)* | -2 (3)* | -41 (5) |
| Hel2 (245-469)... | | LEQVVYKPQKF | 448 | 458 | -3 (5)* | 1 (3)* | -36 (4) |
| | | EQVVYKPQKF | 449 | 458 | -4 (5)* | 0 (3)* | -28 (3) |
| | | FRKVESRISDKFKYIIAQL | 459 | 477 | | -1 (2)* | -20 (2) |
| | | MRDTESLAKRICKDLENL | 478 | 495 | | -2 (2)* | -13 (3) |
| | | AKRICKDLENL | 485 | 495 | -7 (5) | -2 (3)* | -21 (5) |
| | RNA binding region (505-520) | SQIQNREFGTQKYEQW | 496 | 511 | -4 (4)* | 0 (2)* | -43 (4) |
| | | IVTVQKACM | 512 | 520 | -2 (2)* | -7 (2) | -3 (4)* |
| | | IVTVQKACM | 512 | 520 | -1 (5)* | | -1 (2)* |
| | CARD2/CTD :Hel2i latch (520-540) | VFQMPDKDEESRICKA | 521 | 536 | -12 (5) | -4 (3)* | -31 (3) |
| Hel2i (470-607) | | VFQMPDKDEESRICKA | 521 | 536 | -10 (4) | -1 (3)* | -29 (3) |
| | | VFQMPDKDEESRICKALF | 521 | 538 | -8 (4) | -2 (2)* | -22 (3) |
| | | LYTSHLRKYNDAL | 539 | 551 | 0 (1)* | 2 (1)* | 2 (2)* |
| | | DALIISEHARMKD | 549 | 561 | 1 (3)* | 0 (1)* | -6 (2) |
| | CARD2:Hel2i latch (565-580) | HARMKDALD | 556 | 564 | 8 (50) | -1 (1)* | -12 (3) |
| | | HARMKDALD | 556 | 564 | 8 (49) | -1 (1)* | -14 (4) |
| | | YLKDFFSNVRAAGF | 565 | 578 | 10 (3) | -1 (2)* | -25 (3) |
| | | ESVSRDPSNENPKLED | 597 | 612 | 1 (4)* | -3 (2)* | -28 (3) |

Consolidated HDX differential peptide coverage (cont.)

| Domain | Feature | Peptide Sequence | Start Residue | End Residue | Δ HDX: wt RIG-I ± 5'ppp 10bp HP RNA | Δ HDX: apo G731R vs. WT RIG-I | Δ HDX: G731R ± 5'ppp 10bp HP RNA |
|---|---|---|---|---|---|---|---|
| Hel2 (608-743) | RNA binding motifs | VSRDPSNENPKLEDLCF | 599 | 615 | -2 (3)* | -1 (3)* | -24 (2) |
| | | FILQEEYHLNPET | 615 | 627 | 0 (4)* | -2 (2)* | -10 (2) |
| | | ILQEEYHLNPET | 616 | 627 | -1 (2)* | -1 (2)* | -13 (2) |
| | motif IV (630-695) | VKTRALVDALKN | 633 | 644 | -7 (2) | -3 (1)* | -15 (3) |
| | | ALKNWIEGNPKL | 641 | 652 | 1 (3)* | -2 (2)* | -13 (2) |
| | | ALKNWIEGNPKL | 641 | 652 | 1 (3)* | -1 (2)* | -12 (2) |
| | | SFLKPGIL | 653 | 660 | -8 (2) | 1 (2)* | -18 (3) |
| | | TGRGKTNQNTGMTLPAQKCIL | 661 | 681 | -9 (4) | -11 (2) | -81 (3) |
| | | DAFKASGDHNIL | 682 | 693 | -9 (3) | 0 (3)* | -53 (4) |
| | | DAFKASGDHNIL | 682 | 693 | -9 (3) | -5 (2)* | -42 (3) |
| | | FKASGDHNIL | 684 | 693 | -6 (3) | 0 (2)* | -35 (3) |
| | | FKASGDHNIL | 684 | 693 | -6 (3) | 0 (2)* | -34 (2) |
| | motif V (695-720) | IATSVADEGID | 694 | 704 | -14 (3) | -5 (2)* | -41 (5) |
| | | IATSVADEGID | 694 | 704 | -13 (4) | -4 (2)* | -35 (3) |
| | | IAQCNL | 705 | 710 | -7 (4) | -3 (3)* | -40 (3) |
| | | VILYE | 711 | 715 | -1 (1)* | 1 (2)* | -2 (3)* |
| | | YEYVGNVIKM | 714 | 723 | -14 (4) | -4 (3)* | -38 (3) |
| | | FLLTSNA | 738 | 744 | -9 (5) | -6 (1) | -33 (3) |
| Pincer (744-803) | Helicase bridge / CTD linker | LTSNAGVIEKEQINM | 740 | 754 | -3 (4)* | -8 (3) | -19 (4) |
| | | YKEKMMNDSIL | 755 | 765 | 0 (1)* | 5 (2)* | -7 (2) |
| | | QTWDEAVF | 768 | 775 | -1 (3)* | -1 (2)* | -41 (4) |
| | | WDEAVF | 770 | 775 | 1 (2)* | -1 (3)* | -18 (2) |
| CTD (804-925) | | VRVIEECHY | 822 | 830 | -2 (4)* | -2 (1)* | -14 (2) |
| | | TVLGDA | 831 | 836 | 0 (3)* | | -30 (3) |
| | 5' capping loop (840-860) | TVLGDAFKECF | 831 | 841 | -2 (4)* | 0 (2)* | |
| | | VSRPHPKPKQFSSF | 842 | 855 | | -6 (1) | -80 (3) |
| | RNA binding region (880-905) | CARQNCSHDWGIHVKYKTFEIPVIKIESF | 863 | 891 | | -1 (1)* | -9 (3) |
| | | VVEDIATGVQTL | 892 | 903 | -11 (3) | -6 (3) | -30 (3) |
| | | DIATGVQTL | 895 | 903 | -10 (3) | -8 (3) | -28 (6) |
| | | YSKWKDFHF | 904 | 912 | | -6 (2) | -33 (3) |
| | | EKIPFDPAEMSK | 913 | 924 | -2 (3)* | -7 (2) | -38 (5) |
| | | EKIPFDPAEMSK | 913 | 924 | -1 (4)* | -7 (2) | -53 (3) |

**Figure S3. Annotated mapping of consolidated HDX differential peptide coverage related to data in figure 4.**

Chart summarizing the differential HDX patterns of specific peptide of RIG-I from each experimental condition, annotated by protein domain and functional features.

**Figures S4-S9. Hydrogen-Deuterium Exchange Mass Spectroscopy sequence coverage heatmaps related to data in figure 4.**

Time-consolidated HDX data is shown using a perturbation view from HDX Workbench (Scripps Research Institute, 2024). Individual peptides are represented by horizontal strips below their respective portion of the protein sequence with values representing the percent deuterium uptake ± standard deviation. Individual peptides are colored according to the colorimetric perturbation key shown.

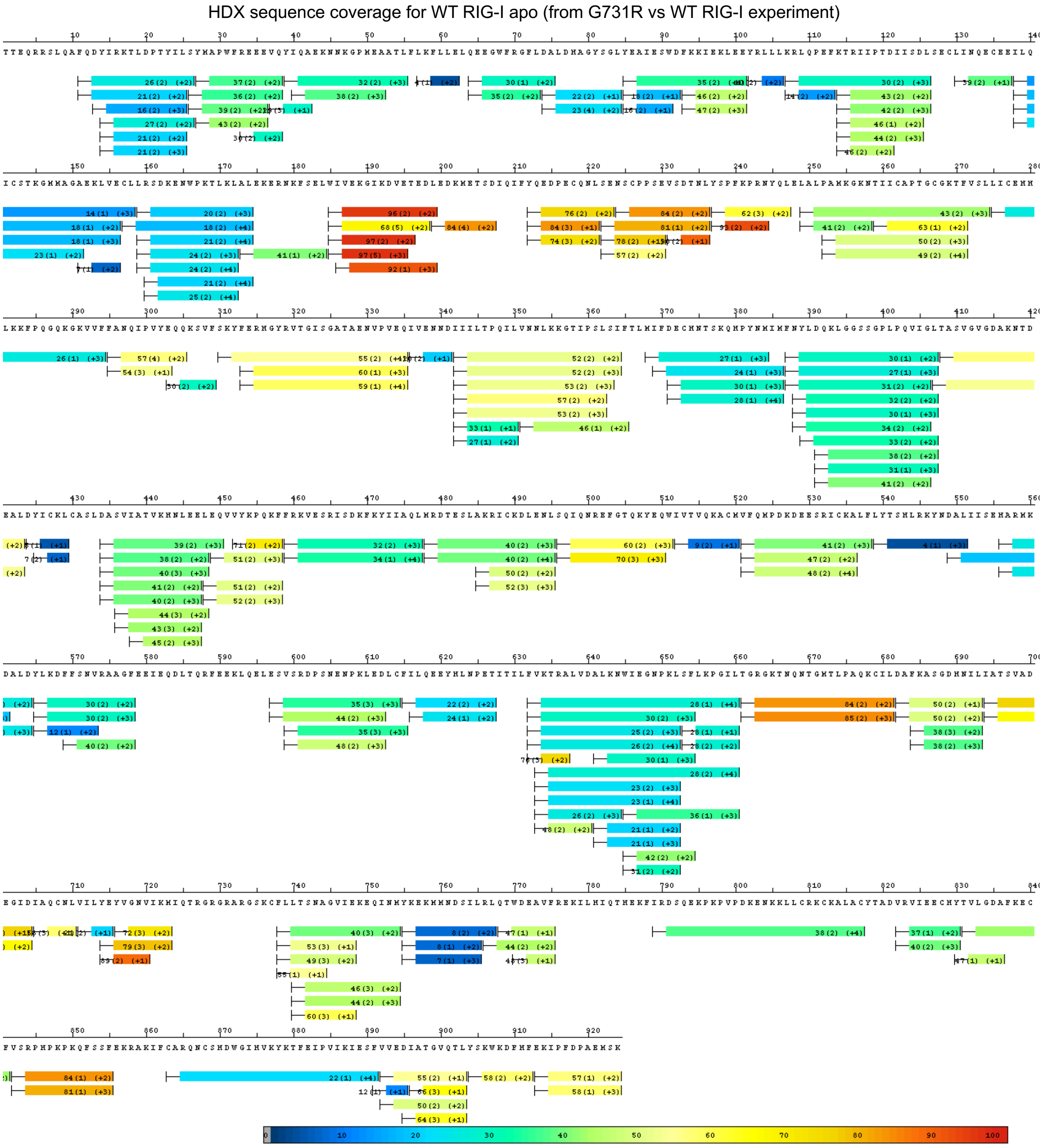

**Figure S4. HDX sequence coverage for WT RIG-I apo (from G731R vs WT RIG-I experiment)**

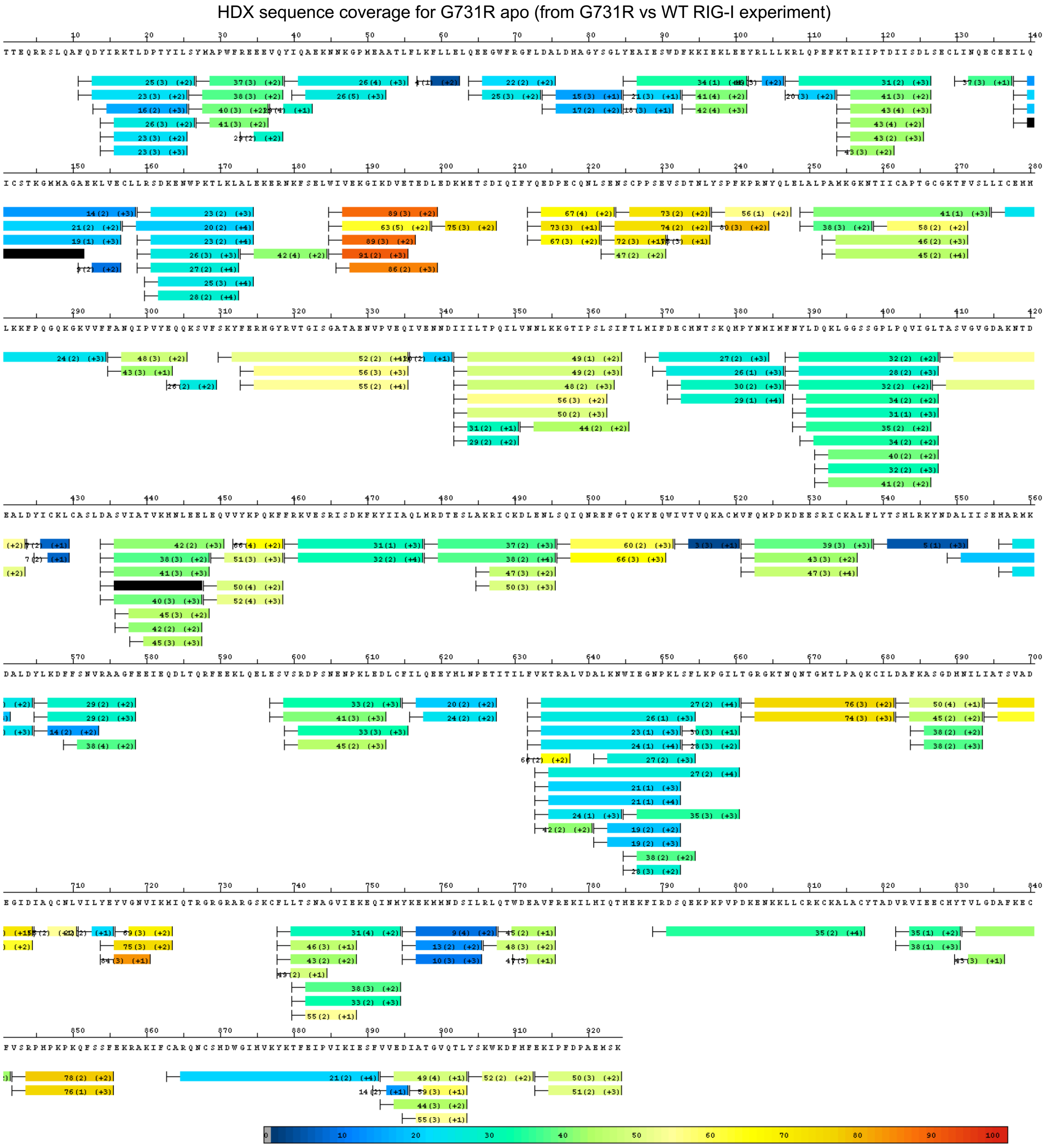


**Figure S5. HDX sequence coverage for G731R apo (from G731R vs WT RIG-I experiment)**

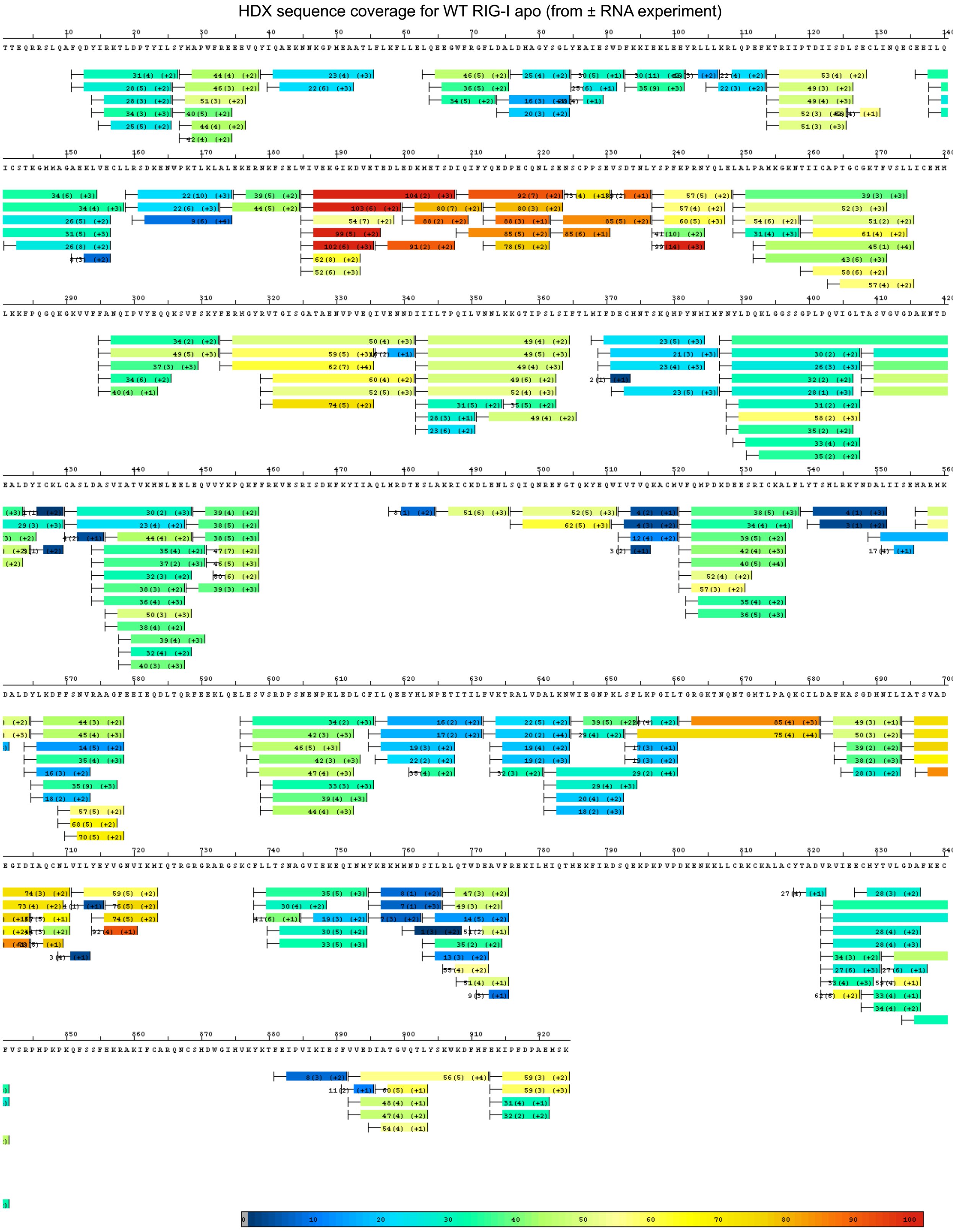


**Figure S6. HDX sequence coverage for WT RIG-I apo (from ± RNA experiment)**

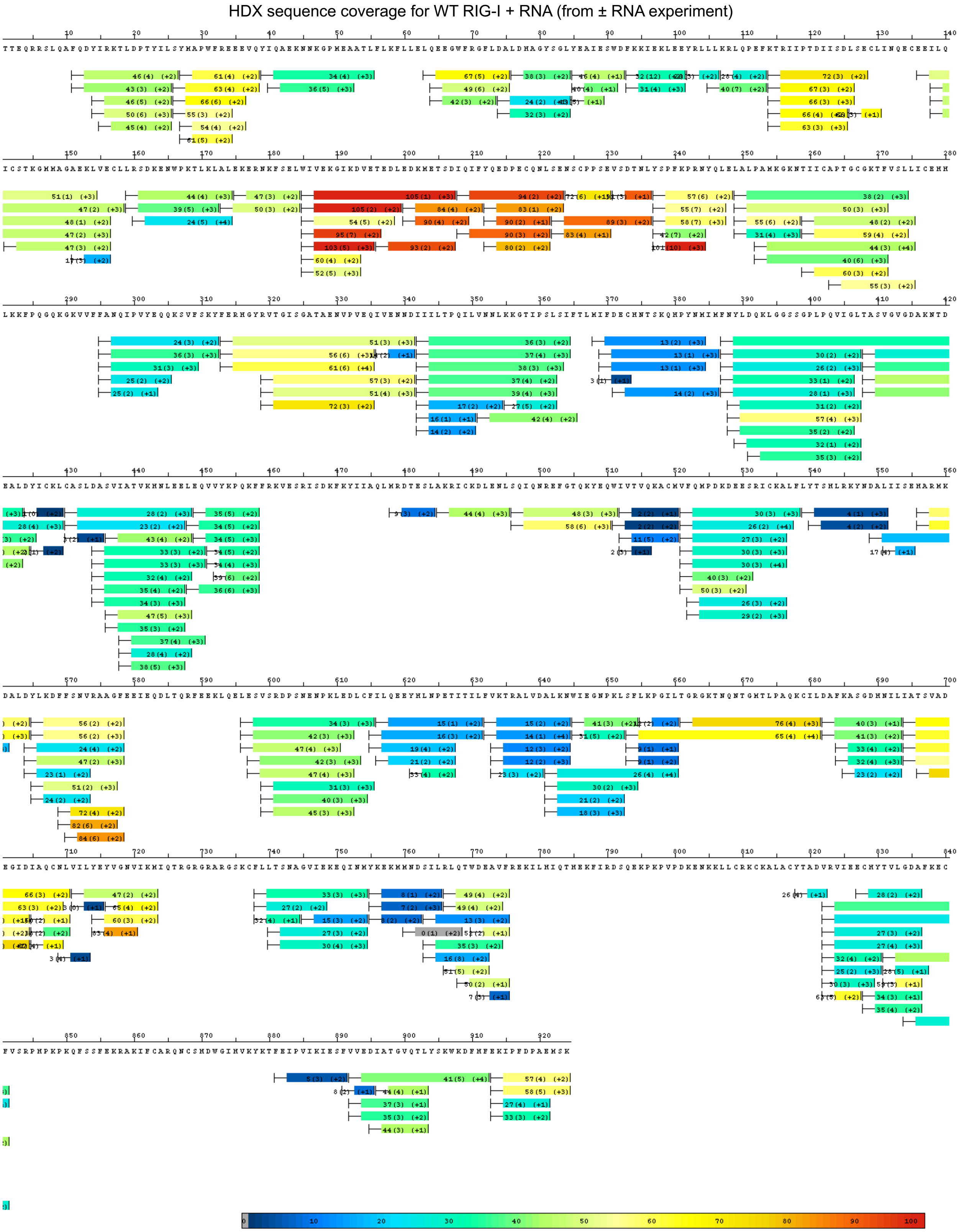


**Figure S7. HDX sequence coverage for WT RIG-I + RNA (from ± RNA experiment)**

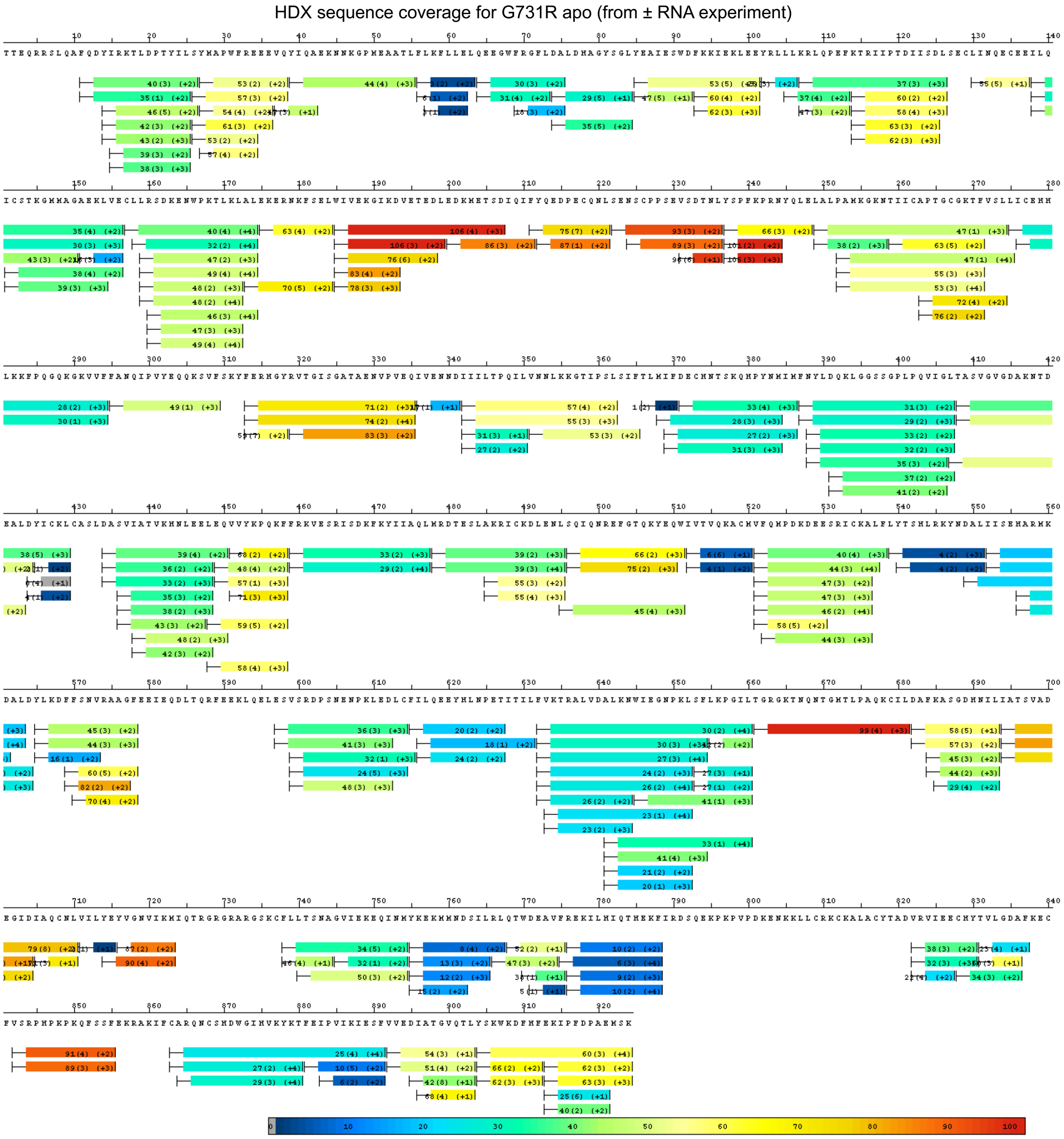

### Figure S8. HDX sequence coverage for G731R apo (from ± RNA experiment)

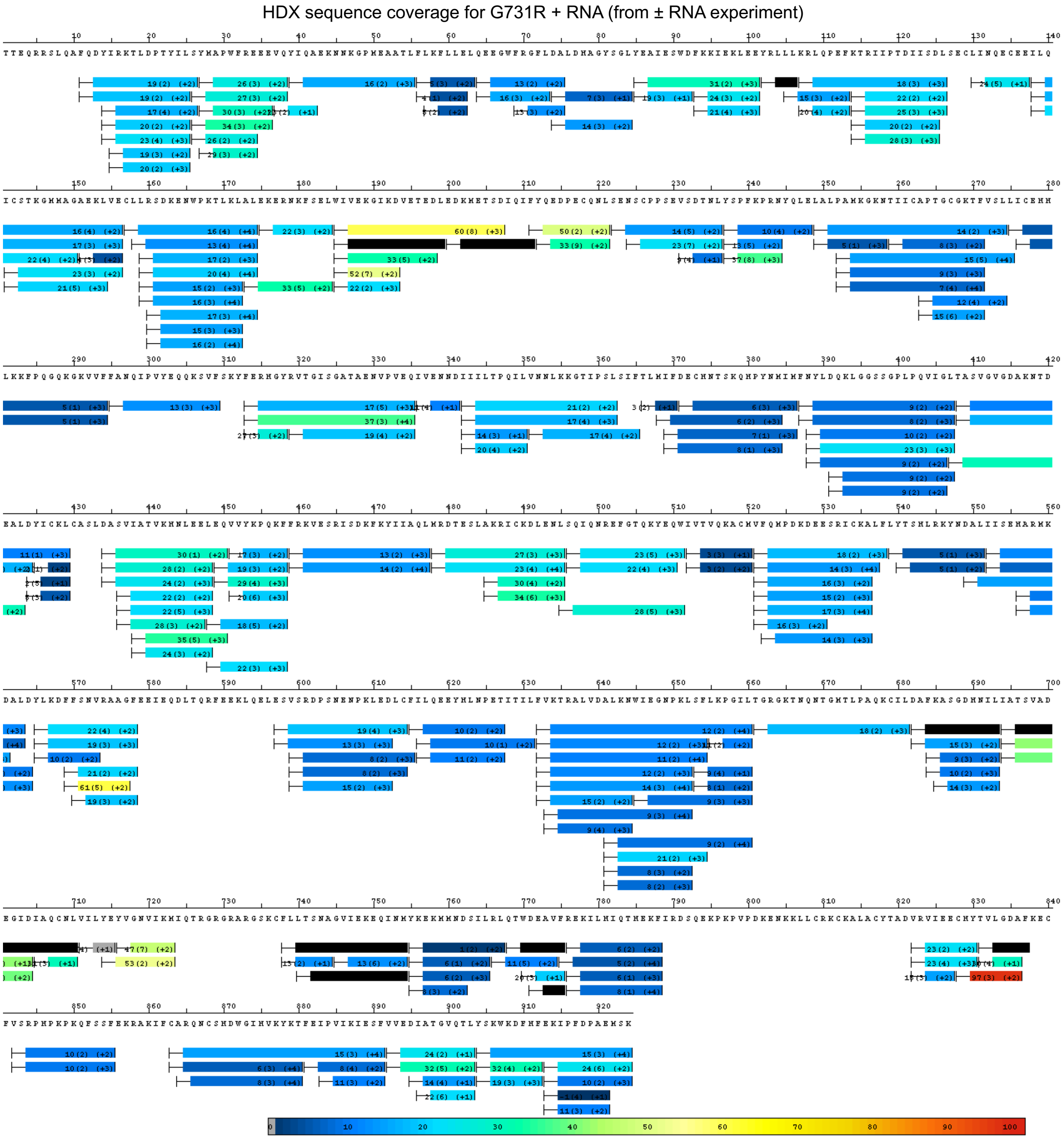


**Figure S9. HDX sequence coverage for G731R + RNA (from ± RNA experiment)**

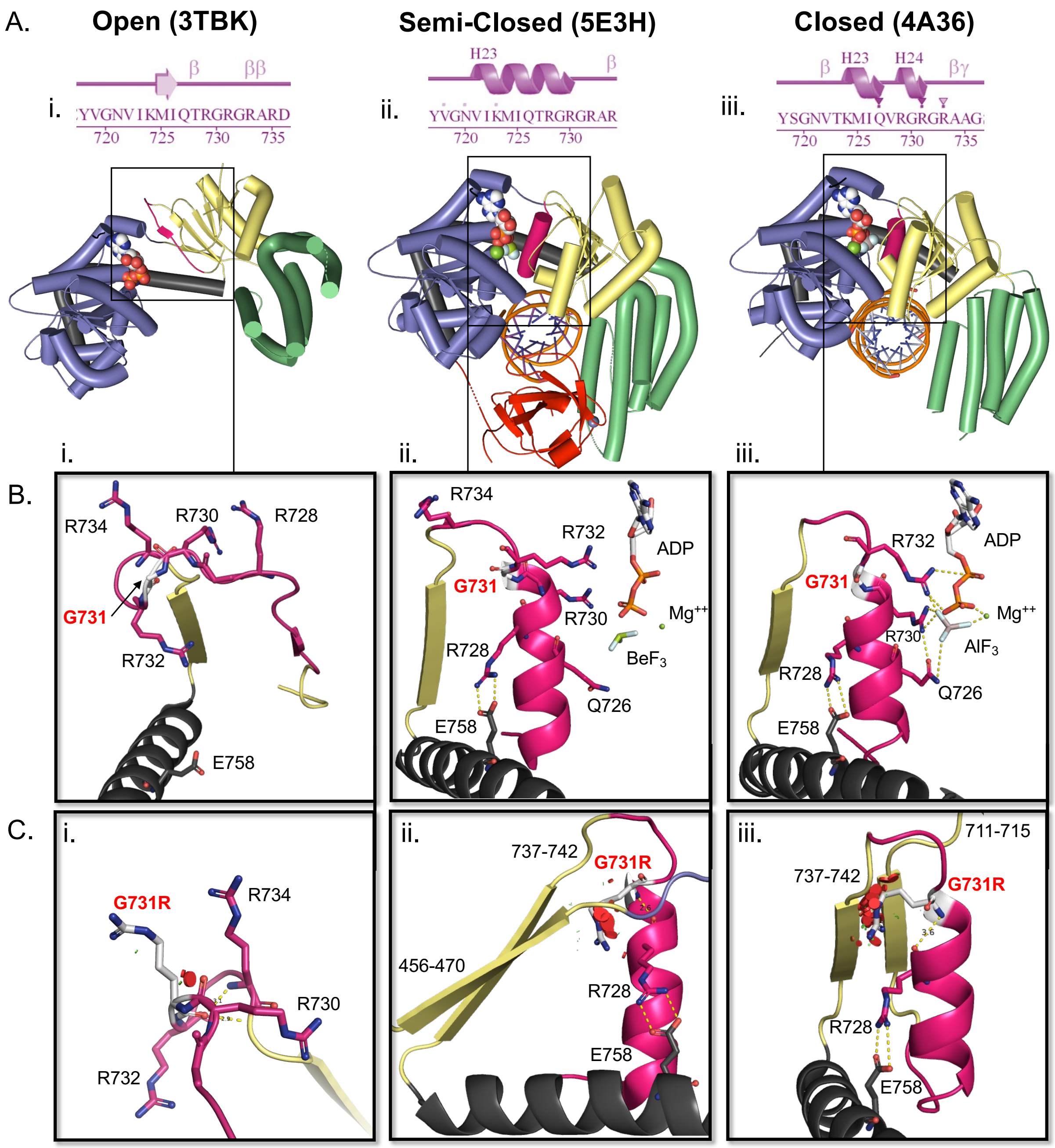


**Figure S10. Conformational changes in RIG-I from open to closed states upon RNA binding**

(A) PDBsum linear representation of secondary structure formation in motif VI (in pink) of (i) open mouse RIG-I helicases (PDB: 3TBK), (ii) semi-closed human RIG-I (PDB: 5E3H), and (iii) closed duck RIG-I helicases (PDB: 4A36).Below are front top views of

RIG-I crystal structures captured in each conformational state. Structures are aligned by the Hel1 domain.

(B) Zoomed in view of each respective RIG-I structure showing the isolated motif VI and associated interactions in detail.

(C) PyMOL Mutagenesis Wizard analysis of G731R computational mutation in each respective structure. The red disks indicate pairwise overlap of atomic van der Waals radii.

**Table S1**: Whole Genome Sequencing analysis summary
(see excel file)

**Table S2**: RNA binding measurements of WT RIG-I and G731R by fluorescence polarization were determined from experiments shown in figures 3B and S2.

| RNA | constant | WT RIG-I | G731R |
|---|---|---|---|
| 5'ppp 10bp HP (FAM) | $K_D$ | 1.2 ± 0.03 nM | 0.83 ± 0.25 nM |
| 5'ppp ds39 | $IC_{50}$ | 27.8 ± 2.4 nM | 15.7 ± 1.4 nM |
| 5'ppp ds39 | $K_i$ | 3.0 ± 0.3 nM | 1.2 ± 0.4 nM |

**Table S3**: RNA unbinding measurements of WT RIG-I and G731R by stopped-flow fluorescence intensity were determined from experiments shown in figure 5 C,D,E.

| | WT | | G731R | | |
|---|---|---|---|---|---|
| | CTD | Helicase | CTD | Helicase | |
| $k_{off}$ ($s^{-1}$) | 0.141 ± 0.011 | 0.0045 ± 0.00006 | 0.147 ±0.015 | 0.0062 ± 0.003 | - ATP |
| Kinetic mode population (%) | 75.1 ± 4.26 | 24.9 ± 4.26 | 92.4 ± 2.03 | 7.6 ± 2.03 | |
| Kinetic mode lifetime (s) | 7.1 ± 0.5 | 223 ± 2.8 | 6.7 ± 0.7 | 198 ± 76.0 | |
| $R^2$ | 0.9829 ± 0.01123 | | 0.9814 ± 0.01063 | | |
| $k_{off}$ ($s^{-1}$) | 0.128 ± 0.012 | 0.0105 ± 0.001 | 0.137 ± 0.010 | 0.0096 ± 0.006 | + ATP |
| Kinetic mode population (%) | 76.7 ± 0.99 | 23.3 ± 0.99 | 95.2 ± 0.74 | 4.8 ± 0.74 | |
| Kinetic mode lifetime (s) | 7.9 ± 0.7 | 96 ± 10.9 | 7.4 ± 0.5 | 176 ± 127 | |
| $R^2$ | 0.9875 ± 0.007793 | | 0.9822 ± 0.007674 | | |

**Table S4**: Comparison of CADD and AlphaMissense scores to biochemical impact, as determined from experiments shown in figure 7A, for selected G731 missense mutants.

| Genomic DNA change | cDNA change | Protein variant | Original | Nucleotide changed | CADD score | AlphaMissense score | AlphaMissense class | MAF in gnomAD | IFNβ reporter activity (relative to WT) |
|---|---|---|---|---|---|---|---|---|---|
| Chr9:32466433C>G | c.2192G>C | p.Gly731Ala | GGA | GCA | 24.2 | 0.308 | likely_benign | 0 | GOF |
| Chr9:32466433C>T | c.2192G>A | p.Gly731Glu | GGA | GAA | 25.7 | 0.97 | likely_pathogenic | 6.24E-07 | LOF |
| Chr9:32466434C>G | c.2191G>C | p.Gly731Arg | GGA | CGA | 25.5 | 0.973 | likely_pathogenic | Novel (this report) | LOF |
| Chr9:32466434C>T | c.2191G>A | p.Gly731Arg | GGA | AGA | 25.7 | 0.973 | likely_pathogenic | 0 | LOF |
| Chr9:32466433C>A | c.2192G>T | p.Gly731Val | GGA | GTA | 25.3 | 0.836 | likely_pathogenic | 0 | Not tested |
| | - | p.Gly731Leu | GGA | CTA | - | 0.926 | likely_pathogenic | - | GOF |
| | - | p.Gly731Phe | GGA | TTT | - | 0.983 | likely_pathogenic | - | Partial LOF |
| | - | p.Gly731Lys | GGA | AAA | - | 0.991 | likely_pathogenic | - | LOF |

MAF: Minor Allele Frequency, CADD: Combined Annotation Dependent Depletion, LOF: Loss Of Function, GOF: Gain Of Function
