## Supplemental Table for "Human RIG-I Antiviral Deficiency Caused by a Dominant-Negative Variant Locked in a Signaling-Inactive State"

### Potential Compound heterozygous model

| gene | biotype | chrom | start | end | ref |
|---|---|---|---|---|---|
| GPR84 | protein_codin | chr12 | 54757430 | 54757431 | A |
| GPR84 | protein_codin | chr12 | 54757552 | 54757554 | AC |
| NBEAL1 | protein_codin | chr2 | 203964368 | 203964369 | G |
| NBEAL1 | protein_codin | chr2 | 203972819 | 203972820 | G |
| CDON | protein_codin | chr11 | 125887164 | 125887165 | G |
| CDON | protein_codin | chr11 | 125831575 | 125831576 | T |
| ARSD | protein_codin | chrX | 2835988 | 2835989 | A |
| ARSD | protein_codin | chrX | 2835994 | 2835995 | C |
| ARSD | protein_codin | chrX | 2836040 | 2836041 | A |
| ARSD | protein_codin | chrX | 2836046 | 2836047 | C |
| CTD-3088G3.8 | protein_codin | chr16 | 11501174 | 11501175 | C |
| CTD-3088G3.8 | protein_codin | chr16 | 11572453 | 11572454 | C |

### Autosomal dominant model

| gene | biotype | chrom | start | end | ref |
|---|---|---|---|---|---|
| UBAP2 | protein_codin | chr9 | 33944385 | 33944386 | G |
| DDX58* | protein_codin | chr9 | 32466433 | 32466434 | C |
| AIMP1 | protein_codin | chr4 | 107252977 | 107252978 | G |
| PANK2 | protein_codin | chr20 | 3888722 | 3888723 | C |
| TGM1 | protein_codin | chr14 | 24729877 | 24729878 | T |
| WDR5 | protein_codin | chr9 | 137007134 | 137007135 | C |
| CYP19A1 | protein_codin | chr15 | 51507380 | 51507381 | T |
| DRG2 | protein_codin | chr17 | 17997260 | 17997261 | C |
| PDS5B | protein_codin | chr13 | 33332328 | 33332329 | A |
| CSMD2 | protein_codin | chr1 | 34090704 | 34090705 | G |
| BOK | protein_codin | chr2 | 242499105 | 242499106 | C |
| C12orf66 | protein_codin | chr12 | 64609581 | 64609582 | C |
| MBD6 | protein_codin | chr12 | 57919565 | 57919566 | T |
| TECPR2 | protein_codin | chr14 | 102891422 | 102891423 | G |
| PIPOX | protein_codin | chr17 | 27381611 | 27381612 | A |
| KIAA1239 | protein_codin | chr4 | 37448444 | 37448445 | C |
| UBALD2 | protein_codin | chr17 | 74266472 | 74266473 | T |
| KIAA1522 | protein_codin | chr1 | 33237891 | 33237892 | T |
| SEL1L3 | protein_codin | chr4 | 25759185 | 25759186 | A |
| TMEM194A | protein_codin | chr12 | 57453806 | 57453807 | G |
| ABCB4 | protein_codin | chr7 | 87053250 | 87053251 | A |
| PKHD1L1 | protein_codin | chr8 | 110499001 | 110499002 | G |
| HEATR1 | protein_codin | chr1 | 236715384 | 236715385 | G |
| A2ML1 | protein_codin | chr12 | 9027042 | 9027043 | A |
| ZNF717 | protein_codin | chr3 | 75786765 | 75786766 | T |

| | | | | | |
|---|---|---|---|---|---|
| PHLPP1 | protein_codin | chr18 | 60383070 | 60383071 | A |
| SLC25A15 | protein_codin | chr13 | 41367414 | 41367415 | C |
| ANKRD6 | protein_codin | chr6 | 90333715 | 90333716 | T |
| OR52M1 | protein_codin | chr11 | 4566991 | 4566992 | C |
| FOXE3 | protein_codin | chr1 | 47882525 | 47882526 | C |
| ZNF292 | protein_codin | chr6 | 87968685 | 87968686 | C |
| WHSC1 | protein_codin | chr4 | 1955152 | 1955153 | A |
| DAPK1 | protein_codin | chr9 | 90283552 | 90283553 | C |
| ARHGEF38 | protein_codin | chr4 | 106580386 | 106580387 | C |
| R3HDML | protein_codin | chr20 | 42966024 | 42966025 | T |
| ZNF736 | protein_codin | chr7 | 63809516 | 63809517 | A |
| TAF15 | protein_codin | chr17 | 34171786 | 34171787 | G |
| THNSL2 | protein_codin | chr2 | 88472861 | 88472862 | T |
| FIGNL1 | protein_codin | chr7 | 50514362 | 50514363 | C |
| CTD-3088G3.8 | protein_codin | chr16 | 11572453 | 11572454 | C |
| AFG3L2 | protein_codin | chr18 | 12377022 | 12377023 | T |
| THAP2 | protein_codin | chr12 | 72058653 | 72058654 | G |
| ZNF318 | protein_codin | chr6 | 43306268 | 43306269 | C |
| HUG1 | protein_codin | chr10 | 102883591 | 102883592 | A |
| PRMT2 | protein_codin | chr21 | 48071771 | 48071772 | C |
| CEACAM6 | protein_codin | chr19 | 42265216 | 42265217 | C |
| ZNF304 | protein_codin | chr19 | 57867583 | 57867584 | T |
| ECSIT | nonsense_me | chr19 | 11621446 | 11621447 | C |
| NRDE2 | protein_codin | chr14 | 90755266 | 90755267 | A |
| CYP21A2 | protein_codin | chr6 | 32007321 | 32007322 | G |
| CUX2 | protein_codin | chr12 | 111537439 | 111537442 | GGT |
| CGNL1 | protein_codin | chr15 | 57839672 | 57839673 | C |
| TARSL2 | protein_codin | chr15 | 102241343 | 102241344 | C |
| PTBP1 | protein_codin | chr19 | 798541 | 798542 | G |
| TOM1L2 | protein_codin | chr17 | 17765687 | 17765688 | C |
| C21orf2 | protein_codin | chr21 | 45750749 | 45750750 | G |
| CCDC116 | protein_codin | chr22 | 21989215 | 21989230 | GCCTGGCTA |
| CHADL | protein_codin | chr22 | 41634042 | 41634044 | CG |
| FOXD4 | protein_codin | chr9 | 117168 | 117170 | AT |
| ICA1L | protein_codin | chr2 | 203653630 | 203653633 | CAG |
| KRTAP4-9 | protein_codin | chr17 | 39261807 | 39261857 | GTCTGTGTG |
| NLRP14 | protein_codin | chr11 | 7078931 | 7078933 | AT |
| PCSK5 | protein_codin | chr9 | 78790206 | 78790207 | C |
| RNF25 | protein_codin | chr2 | 219528797 | 219528799 | TC |
| ATN1 | protein_codin | chr12 | 7045890 | 7045894 | ACAG |
| C16orf3 | protein_codin | chr16 | 90095597 | 90095598 | G |

| | | | | | |
|---|---|---|---|---|---|
| CGNL1 | protein_codin | chr15 | 57839674 | 57839675 | C |
| HRCT1 | protein_codin | chr9 | 35906594 | 35906595 | A |
| KRT10 | protein_codin | chr17 | 38975102 | 38975103 | A |
| RP1L1 | protein_codin | chr8 | 10467636 | 10467637 | T |

summarized WGS analysis for the patient based on those additional filters:

All genetic models: genotyping Quality score > 95

Recessive models: variants without no homozygous individuals in gnomAD database

Dominant models: variants are novel and not present in gnomAD database

No other IFN pathway variant found in the patient other than the RIG-I variant. No homozygous

GEMINI pipeline used for this analysis  is using a different isoform (ENST00000379868.1) for R

| alt | impact | impact_severi | rs_ids | aaf_gnomad_ | gnomad_num | gnomad_num |
|---|---|---|---|---|---|---|
| G | missense_var | MED | rs949956011 | 3.2329E-05 | -1 | -1 |
| A | frameshift_va | HIGH | rs759103894 | 3.2369E-05 | 0 | 15 |
| T | missense_var | MED | rs201657702 | 0.0002268 | 0 | 35 |
| A | missense_var | MED | rs151000588 | None | 0 | 2 |
| A | missense_var | MED | rs138087778 | 0.00029064 | 0 | 78 |
| A | missense_var | MED | rs377706219 | 3.2285E-05 | 0 | 15 |
| C | missense_var | MED | rs143238998 | 0.00035694 | 0 | 1 |
| A | missense_var | MED | rs150899882 | 0.00035681 | 0 | 1 |
| T | missense_var | MED | rs67272620 | 0.00042906 | 0 | 9 |
| T | missense_var | MED | rs67359049 | 0.00042915 | 0 | 8 |
| T | missense_var | MED | rs184717665 | 0.00353019 | 0 | 39 |
| G | missense_var | MED | None | None | -1 | -1 |

| alt | impact | impact_severi | rs_ids | aaf_gnomad_ | gnomad_num | gnomad_num |
|---|---|---|---|---|---|---|
| A | missense_var | MED | rs146320733 | None | -1 | -1 |
| G | missense_var | MED | None | None | -1 | -1 |
| T | missense_var | MED | None | None | -1 | -1 |
| G | missense_var | MED | None | None | -1 | -1 |
| C | missense_var | MED | None | None | -1 | -1 |
| T | missense_var | MED | None | None | -1 | -1 |
| C | missense_var | MED | None | None | -1 | -1 |
| G | missense_var | MED | None | None | -1 | -1 |
| G | missense_var | MED | None | None | -1 | -1 |
| A | missense_var | MED | None | None | -1 | -1 |
| A | missense_var | MED | None | None | -1 | -1 |
| A | missense_var | MED | None | None | -1 | -1 |
| A | missense_var | MED | None | None | -1 | -1 |
| C | missense_var | MED | None | None | -1 | -1 |
| G | missense_var | MED | None | None | -1 | -1 |
| G | missense_var | MED | None | None | -1 | -1 |
| A | missense_var | MED | None | None | -1 | -1 |
| A | missense_var | MED | None | None | -1 | -1 |
| G | missense_var | MED | None | None | -1 | -1 |
| A | missense_var | MED | rs100658318 | None | -1 | -1 |
| T | missense_var | MED | None | None | -1 | -1 |
| T | missense_var | MED | None | None | -1 | -1 |
| C | missense_var | MED | None | None | -1 | -1 |
| C | missense_var | MED | None | None | -1 | -1 |
| G | missense_var | MED | None | None | -1 | -1 |

| | | | | | | |
|---|---|---|---|---|---|---|
| G | missense_var | MED | rs11152356 | None | -1 | -1 |
| T | missense_var | MED | None | None | -1 | -1 |
| A | missense_var | MED | None | None | -1 | -1 |
| A | missense_var | MED | None | None | -1 | -1 |
| G | missense_var | MED | None | None | -1 | -1 |
| T | missense_var | MED | None | None | -1 | -1 |
| G | missense_var | MED | None | None | -1 | -1 |
| G | missense_var | MED | None | None | -1 | -1 |
| A | missense_var | MED | None | None | -1 | -1 |
| G | missense_var | MED | None | None | -1 | -1 |
| C | missense_var | MED | None | None | -1 | -1 |
| A | missense_var | MED | None | None | -1 | -1 |
| C | missense_var | MED | None | None | -1 | -1 |
| T | missense_var | MED | None | None | -1 | -1 |
| G | missense_var | MED | None | None | -1 | -1 |
| A | missense_var | MED | None | None | -1 | -1 |
| T | missense_var | MED | None | None | -1 | -1 |
| T | missense_var | MED | None | None | -1 | -1 |
| G | missense_var | MED | rs937838297 | None | -1 | -1 |
| G | missense_var | MED | None | None | -1 | -1 |
| T | missense_var | MED | None | None | -1 | -1 |
| A | missense_var | MED | None | None | -1 | -1 |
| T | missense_var | MED | rs931413131 | None | -1 | -1 |
| G | missense_var | MED | None | None | -1 | -1 |
| A | splice_accept | HIGH | None | None | -1 | -1 |
| G | splice_donor_ | HIGH | None | None | -1 | -1 |
| CTGAGT | stop_gained | HIGH | None | None | -1 | -1 |
| T | missense_var | MED | rs143491081 | None | 0 | 0 |
| A | missense_var | MED | rs568643597 | None | 0 | 0 |
| T | missense_var | MED | rs962451906 | None | 0 | 0 |
| A | missense_var | MED | rs749720437 | None | -1 | -1 |
| G | frameshift_va | HIGH | None | None | -1 | -1 |
| C | frameshift_va | HIGH | None | None | -1 | -1 |
| A | frameshift_va | HIGH | None | None | -1 | -1 |
| C | frameshift_va | HIGH | None | None | -1 | -1 |
| G | frameshift_va | HIGH | None | None | -1 | -1 |
| A | frameshift_va | HIGH | None | None | -1 | -1 |
| CGAATA | frameshift_va | HIGH | rs71372053,r | None | -1 | -1 |
| T | frameshift_va | HIGH | None | None | -1 | -1 |
| A | inframe_delet | MED | None | None | -1 | -1 |
| GGGGCAGC( | inframe_inser | MED | None | None | -1 | -1 |

| | | | | | | | |
|---|---|---|---|---|---|---|---|
| CTGA | inframe_inser | MED | None | None | | -1 | -1 |
| ACCACCCCC | inframe_inser | MED | rs762598108 | None | | -1 | -1 |
| AGCTGCCGC | inframe_inser | MED | None | None | | -1 | -1 |
| TCCTCTAACT | inframe_inser | MED | rs369606728 | None | | -1 | -1 |

s candidate for the patient.

IG-I so in the list, the variant is listed as p.Gly528Arg instead of p.Gly731Arg.

| gnomad_num | is_coding | codon_chang | aa_change | aa_length | exon | vep_hgvsc |
|---|---|---|---|---|---|---|
| -1 | 1 | Tac/Cac | Y/H | 69/396 | 2/2 | ENST0000026 |
| 250466 | 1 | Gtg/tg | V/X | 28/396 | 2/2 | ENST0000026 |
| 156036 | 1 | ttG/ttT | L/F | 372/2694 | 11/55 | ENST0000044 |
| 157674 | 1 | Gcc/Acc | A/T | 591/2694 | 13/55 | ENST0000044 |
| 251288 | 1 | cCg/cTg | P/L | 249/1264 | 6/20 | ENST0000026 |
| 249114 | 1 | cAg/cTg | Q/L | 1225/1287 | 19/20 | ENST0000039 |
| 123485 | 1 | tTt/tGt | F/C | 240/593 | 5/10 | ENST0000038 |
| 123661 | 1 | tGc/tTc | C/F | 238/593 | 5/10 | ENST0000038 |
| 118743 | 1 | Ttc/Atc | F/I | 223/593 | 5/10 | ENST0000038 |
| 117299 | 1 | Ggt/Agt | G/S | 221/593 | 5/10 | ENST0000038 |
| 6206 | 1 | Gag/Aag | E/K | 2236/2491 | 45/50 | ENST0000059 |
| -1 | 1 | Ggg/Cgg | G/R | 731/2491 | 16/50 | ENST0000059 |

| gnomad_num | is_coding | codon_chang | aa_change | aa_length | exon | vep_hgvsc |
|---|---|---|---|---|---|---|
| -1 | 1 | Cgg/Tgg | R/W | 508/1119 | 14/29 | ENST0000036 |
| -1 | 1 | Gga/Cga | G/R | 528/722 | 15/17 | ENST0000037 |
| -1 | 1 | Gtc/Ttc | V/F | 181/312 | 5/7 | ENST0000035 |
| -1 | 1 | tCt/tGt | S/C | 260/570 | 2/7 | ENST0000031 |
| -1 | 1 | Acg/Gcg | T/A | 179/817 | 4/15 | ENST0000020 |
| -1 | 1 | aCc/aTc | T/I | 110/334 | 5/14 | ENST0000035 |
| -1 | 1 | Atg/Gtg | M/V | 303/503 | 8/10 | ENST0000026 |
| -1 | 1 | Ctg/Gtg | L/V | 67/364 | 2/13 | ENST0000022 |
| -1 | 1 | cAa/cGa | Q/R | 1054/1447 | 27/35 | ENST0000031 |
| -1 | 1 | Ctc/Ttc | L/F | 689/1167 | 13/24 | ENST0000037 |
| -1 | 1 | Ctg/Atg | L/M | 70/212 | 2/5 | ENST0000031 |
| -1 | 1 | Gtt/Ttt | V/F | 133/468 | 2/4 | ENST0000031 |
| -1 | 1 | cTg/cAg | L/Q | 272/1003 | 6/13 | ENST0000035 |
| -1 | 1 | gGg/gCg | G/A | 249/1411 | 6/20 | ENST0000035 |
| -1 | 1 | tAt/tGt | Y/C | 237/390 | 5/8 | ENST0000032 |
| -1 | 1 | gCt/gGt | A/G | 1612/1742 | 7/7 | ENST0000030 |
| -1 | 1 | Tcc/Acc | S/T | 128/164 | 3/3 | ENST0000032 |
| -1 | 1 | Tcc/Acc | S/T | 979/1035 | 6/7 | ENST0000037 |
| -1 | 1 | Tgg/Cgg | W/R | 1042/1097 | 23/24 | ENST0000026 |
| -1 | 1 | aCg/aTg | T/M | 397/444 | 9/9 | ENST0000030 |
| -1 | 1 | Ttt/Att | F/I | 728/1286 | 17/28 | ENST0000026 |
| -1 | 1 | Gca/Tca | A/S | 3278/4243 | 59/78 | ENST0000037 |
| -1 | 1 | aCt/aGt | T/S | 2006/2063 | 43/44 | ENST0000036 |
| -1 | 1 | tAc/tCc | Y/S | 1415/1454 | 34/36 | ENST0000029 |
| -1 | 1 | Acg/Ccg | T/P | 663/907 | 5/5 | ENST0000040 |

| | | | | | | |
|---|---|---|---|---|---|---|
| -1 | 1 | gAg/gGg | E/G | 52/1717 | 1/17 | ENST0000026 |
| -1 | 1 | gCa/gTa | A/V | 18/301 | 2/7 | ENST0000033 |
| -1 | 1 | caT/caA | H/Q | 386/727 | 12/16 | ENST0000033 |
| -1 | 1 | aCa/aAa | T/K | 191/317 | 1/1 | ENST0000036 |
| -1 | 1 | cCa/cGa | P/R | 180/319 | 1/1 | ENST0000033 |
| -1 | 1 | gCt/gTt | A/V | 1775/2718 | 8/8 | ENST0000033 |
| -1 | 1 | tAc/tGc | Y/C | 95/713 | 2/12 | ENST0000038 |
| -1 | 1 | caC/caG | H/Q | 655/1430 | 19/26 | ENST0000035 |
| -1 | 1 | aaC/aaA | N/K | 470/777 | 10/14 | ENST0000042 |
| -1 | 1 | agT/agG | S/R | 76/253 | 1/5 | ENST0000021 |
| -1 | 1 | Aag/Cag | K/Q | 426/427 | 5/5 | ENST0000035 |
| -1 | 1 | gGt/gAt | G/D | 492/589 | 15/16 | ENST0000031 |
| -1 | 1 | Tct/Cct | S/P | 65/484 | 1/8 | ENST0000032 |
| -1 | 1 | gGt/gAt | G/D | 208/674 | 4/4 | ENST0000035 |
| -1 | 1 | Ggg/Cgg | G/R | 731/2491 | 16/50 | ENST0000059 |
| -1 | 1 | cAg/cTg | Q/L | 20/797 | 1/17 | ENST0000026 |
| -1 | 1 | gGc/gTc | G/V | 26/52 | 2/2 | ENST0000054 |
| -1 | 1 | Gta/Ata | V/I | 1823/2279 | 10/10 | ENST0000036 |
| -1 | 1 | Agc/Ggc | S/G | 353/362 | 1/1 | ENST0000059 |
| -1 | 1 | Ctt/Gtt | L/V | 237/284 | 7/7 | ENST0000033 |
| -1 | 1 | gCt/gTt | A/V | 162/344 | 3/6 | ENST0000019 |
| -1 | 1 | cTg/cAg | L/Q | 116/659 | 3/3 | ENST0000028 |
| -1 | 0 | cGt/cAt | R/H | 48/71 | 2/6 | ENST0000059 |
| -1 | 1 | Tct/Cct | S/P | 818/1164 | 11/14 | ENST0000035 |
| -1 | 0 | | | | | ENST0000041 |
| -1 | 0 | | | | | ENST0000039 |
| -1 | 1 | -/TGAGT | -/* | 1298-1299/13 | 19/19 | ENST0000028 |
| 237622 | 1 | tGt/tAt | C/Y | 422/802 | 10/19 | ENST0000033 |
| 10 | 1 | gGc/gAc | G/D | 4/56 | 1/4 | ENST0000058 |
| 3522 | 1 | tGc/tAc | C/Y | 35/39 | 2/2 | ENST0000046 |
| -1 | 1 | Cct/Tct | P/S | 199/255 | 6/7 | ENST0000032 |
| -1 | 1 | CCTGGCTACT | PGYCP/X | 289-293/613 | 4/5 | ENST0000029 |
| -1 | 1 | ggC/gg | G/X | 344/762 | 3/6 | ENST0000021 |
| -1 | 1 | gAt/gt | D/X | 317/439 | 1/1 | ENST0000038 |
| -1 | 1 | tCT/t | S/X | 388/482 | 11/13 | ENST0000035 |
| -1 | 1 | TCTGTGTGCT | SVCCQPTCSF | 57-73/210 | 1/1 | ENST0000039 |
| -1 | 1 | Ttt/tt | F/X | 773/1093 | 7/12 | ENST0000029 |
| -1 | 1 | cga/cGAATAg | R/RIX | 688/690 | 14/14 | ENST0000037 |
| -1 | 1 | Gag/ag | E/X | 421/459 | 10/10 | ENST0000029 |
| -1 | 1 | CAG/- | Q/- | 488/1190 | 5/10 | ENST0000035 |
| -1 | 1 | ccc/ccTGCAG | P/PAACPVGC | 51/117 | 1/1 | ENST0000040 |

| | | | | | | |
|---|---|---|---|---|---|---|
| -1 | 1 | gcc/gcTGAc | A/AD | 1299/1302 | 19/19 | ENST0000028 |
| -1 | 1 | cac/caCCAC | H/HHPHR | 104/115 | 1/1 | ENST0000035 |
| -1 | 1 | -/AGCTCCGG | -/SSGGGYGG | 561-562/584 | 7/8 | ENST0000026 |
| -1 | 1 | gaa/gGGACT | E/GTKVIEGLQ | 1324/2400 | 4/4 | ENST0000038 |

| vep_hgvsp | cadd_raw | cadd_scaled |
|---|---|---|
| ENSP0000026 | 3.67 | 25.7 |
| ENSP0000026 | None | None |
| ENSP0000039 | 3.07 | 23.8 |
| ENSP0000039 | 3.69 | 25.9 |
| ENSP0000026 | 2.83 | 23.3 |
| ENSP0000037 | 1.66 | 16.59 |
| ENSP0000037 | 1.52 | 15.9 |
| ENSP0000037 | 0.1 | 4.3 |
| ENSP0000037 | 0.08 | 3.89 |
| ENSP0000037 | 1.12 | 14.02 |
| ENSP0000047 | 1.33 | 15.04 |
| ENSP0000047 | 0.63 | 10.5 |

| vep_hgvsp | cadd_raw | cadd_scaled |
|---|---|---|
| ENSP0000035 | 4.27 | 32 |
| ENSP0000036 | 4.14 | 29.5 |
| ENSP0000035 | 4.02 | 28.1 |
| ENSP0000031 | 3.86 | 26.8 |
| ENSP0000020 | 3.84 | 26.7 |
| ENSP0000035 | 3.83 | 26.6 |
| ENSP0000026 | 3.51 | 25.1 |
| ENSP0000022 | 3.43 | 24.8 |
| ENSP0000031 | 3.34 | 24.6 |
| ENSP0000036 | 3.29 | 24.4 |
| ENSP0000031 | 3.28 | 24.3 |
| ENSP0000031 | 3.1 | 23.9 |
| ENSP0000034 | 3.13 | 23.9 |
| ENSP0000035 | 3.08 | 23.8 |
| ENSP0000031 | 3.05 | 23.7 |
| ENSP0000030 | 2.95 | 23.5 |
| ENSP0000033 | 2.83 | 23.3 |
| ENSP0000036 | 2.75 | 23.1 |
| ENSP0000026 | 2.73 | 23.1 |
| ENSP0000030 | 2.77 | 23.1 |
| ENSP0000026 | 2.6 | 22.8 |
| ENSP0000036 | 2.58 | 22.7 |
| ENSP0000035 | 2.46 | 22.5 |
| ENSP0000029 | 2.37 | 22.2 |
| ENSP0000038 | 2.15 | 20.8 |

| | | |
|---|---|---|
| ENSP0000002 | 2.14 | 20.7 |
| ENSP0000003 | 1.96 | 18.8 |
| ENSP0000003 | 1.93 | 18.52 |
| ENSP0000003 | 1.89 | 18.18 |
| ENSP0000003 | 1.58 | 16.17 |
| ENSP0000003 | 1.54 | 15.97 |
| ENSP0000003 | 1.21 | 14.48 |
| ENSP0000003 | 1.12 | 14.01 |
| ENSP0000004 | 1.1 | 13.87 |
| ENSP0000002 | 1.08 | 13.75 |
| ENSP0000003 | 0.96 | 12.99 |
| ENSP0000003 | 0.89 | 12.53 |
| ENSP0000003 | 0.84 | 12.16 |
| ENSP0000003 | 0.75 | 11.45 |
| ENSP0000004 | 0.63 | 10.5 |
| ENSP0000002 | 0.56 | 9.9 |
| ENSP0000004 | 0.19 | 5.7 |
| ENSP0000003 | 0.11 | 4.45 |
| ENSP0000004 | -0.03 | 2.15 |
| ENSP0000003 | -0.08 | 1.58 |
| ENSP0000001 | -0.12 | 1.2 |
| ENSP0000002 | -0.34 | 0.27 |
| ENSP0000004 | -0.37 | 0.23 |
| ENSP0000003 | -0.51 | 0.08 |
| 18967.2:c.550 | 3.37 | 24.6 |
| 97643.3:c.55+ | None | None |
| ENSP0000002 | None | None |
| ENSP0000003 | 4.05 | 28.5 |
| ENSP0000004 | 0.37 | 8.01 |
| ENSP0000004 | 0.2 | 5.85 |
| ENSP0000003 | 0.34 | 7.71 |
| ENSP0000002 | None | None |
| ENSP0000002 | None | None |
| ENSP0000003 | None | None |
| ENSP0000003 | None | None |
| ENSP0000003 | None | None |
| ENSP0000002 | None | None |
| 76767.3:c.*8_ | None | None |
| ENSP0000002 | None | None |
| ENSP0000003 | None | None |
| ENSP0000003 | None | None |

| | | |
|---|---|---|
| ENSP0000028[illegible] | None | None |
| ENSP0000034[illegible] | None | None |
| ENSP0000026[illegible] | None | None |
| ENSP0000037[illegible] | None | None |
